# Automated Skill Optimisation for False Presupposition Handling in Cancer Communication: SkillOpt Versus Conventional Prompt Engineering Across Language Models

**DOI:** 10.64898/2026.09.08.26362559

**Authors:** Sanjay Khanna

**Author notes:** Correspondence: Sanjay Khanna.

## Abstract

**Background:** Large language models (LLMs) may answer cancer questions plausibly while accepting a false presupposition (FP). Prompts that increase FP correction may also challenge questions in which no FP is present (NFP). We compared SkillOpt, an automated skill-optimisation method developed by Microsoft Research, with conventional prompt engineering across models.

**Methods:** Using the physician-verified Cancer-Myth benchmark, GPT-5.4 optimised a written skill for GPT-4o in three independent 12-step executions. Training used 117 FP questions. Rewrites were selected by combined accuracy on a validation set of 58 FP and 30 NFP questions, then tested on 410 unseen FP questions. A separate GPT-4o judged responses. Across seven LLMs, we compared no correction instruction, a deliberately strong seven-example few-shot prompt, three GPT-5.4-generated instruction-only prompts, and full and compact SkillOpt skills. Further tests examined portability, new questions and valid clinical assumptions.

**Results:** SkillOpt increased GPT-4o FP correction from 15.4–17.6% to 82.2–90.5%, gains of 66.3–72.9 percentage points (95% bootstrap confidence intervals 61.7–77.3; all Holm-adjusted p<0.001). Only two or three rewrites were accepted per execution. The first full skill achieved 92.7–96.6% correction on three recipient models but 6.3–16.3% on two weaker models, where few-shot prompting achieved 44.6–54.6%. On Gemini 3.5 Flash and GPT-5.6 Luna, compact SkillOpt retained 90.0–92.9% correction while increasing NFP accuracy from 59.2–79.2% with the full skill to 85.8–93.3%. Both approaches sometimes challenged valid clinical assumptions.

**Conclusions:** SkillOpt produced large, repeatable gains, but it was neither universally portable nor free of over-correction. Automated and conventional prompts should be compared for each intended model using tests of both FP correction and NFP accuracy. Compact SkillOpt may improve this balance on capable models.

## Introduction

A false presupposition (FP) is an incorrect assumption embedded in a question. A large language model (LLM) can answer the requested part accurately while leaving that assumption unchallenged, producing a fluent response that still reinforces misinformation. This differs from hallucination: models may possess the facts needed to reject a false premise but fail to apply them because the question presents the premise as given [1]. In cancer communication, that failure may distort how a patient understands diagnosis, prognosis, treatment or risk.

The problem is established across general and medical question answering. The CREPE benchmark, Open-Domain Question Answering with False Presuppositions, found false presuppositions in 25% of naturally occurring information-seeking questions and showed that systems struggled particularly to determine whether the embedded assumption was factually correct [2]. Health-specific evaluations have reported the same tendency to accept false claims framed as questions [3]. The behaviour is consistent with linguistic accommodation, in which an interlocutor accepts background information introduced by another speaker, and model sycophancy, in which agreement with a user’s framing is favoured over correction [4,5].

Cancer-Myth made this problem measurable in cancer communication. Three haematology-oncology physicians established ground truth for 585 cancer questions containing an FP and 150 controls containing no FP [6]. Across 17 frontier LLMs, no model corrected more than 43% of FPs. Genetic-Pareto (GEPA), a reflective prompt optimiser tested in that study, increased correction to approximately 80%, but incorrectly challenged 41% of NFP controls and reduced performance on other medical benchmarks. Correction and restraint must therefore be assessed together.

Conventional prompt engineering can use explicit rules, worked examples or longer instructions, but performance is sensitive to wording and often changes between models [7]. Automated prompt optimisation makes revision systematic. Optimization by PROmpting (OPRO) generates instructions from previous candidates and scores, while GEPA reflects on execution traces to propose and combine improvements [8,9]. These methods optimise what is measured; they do not ensure that a competing failure such as over-correction is represented adequately during selection.

SkillOpt, developed by Microsoft Research, treats a written skill as the object being trained. A separate optimiser reviews scored responses, proposes bounded revisions and retains a candidate only when held-out validation performance improves, without changing the target model’s weights [10]. It was originally evaluated on non-clinical agentic and reasoning tasks. We tested whether it could optimise a clinical communication behaviour that requires deciding whether correction is warranted, and examined repeatability, learning dynamics, optimiser-target combinations, skill content, portability, conventional prompts, compression, generalisation and over-correction.

## Methods

### Study design and datasets

Cancer-Myth is an expert-reviewed benchmark comprising 585 FP questions and 150 NFP controls. Three haematology-oncology physicians established the ground truth [6]. Using seed 42, we divided FP questions 2:1:7 into 117 training, 58 validation and 410 test questions. Thirty NFP controls were included in validation, leaving 120 reserved controls for later evaluation. FP correction required a response to identify and address the false assumption. NFP accuracy required the model to answer without inventing an FP. Over-correction was the proportion of NFP controls in which the model incorrectly challenged a valid premise.

**Figure 1.**
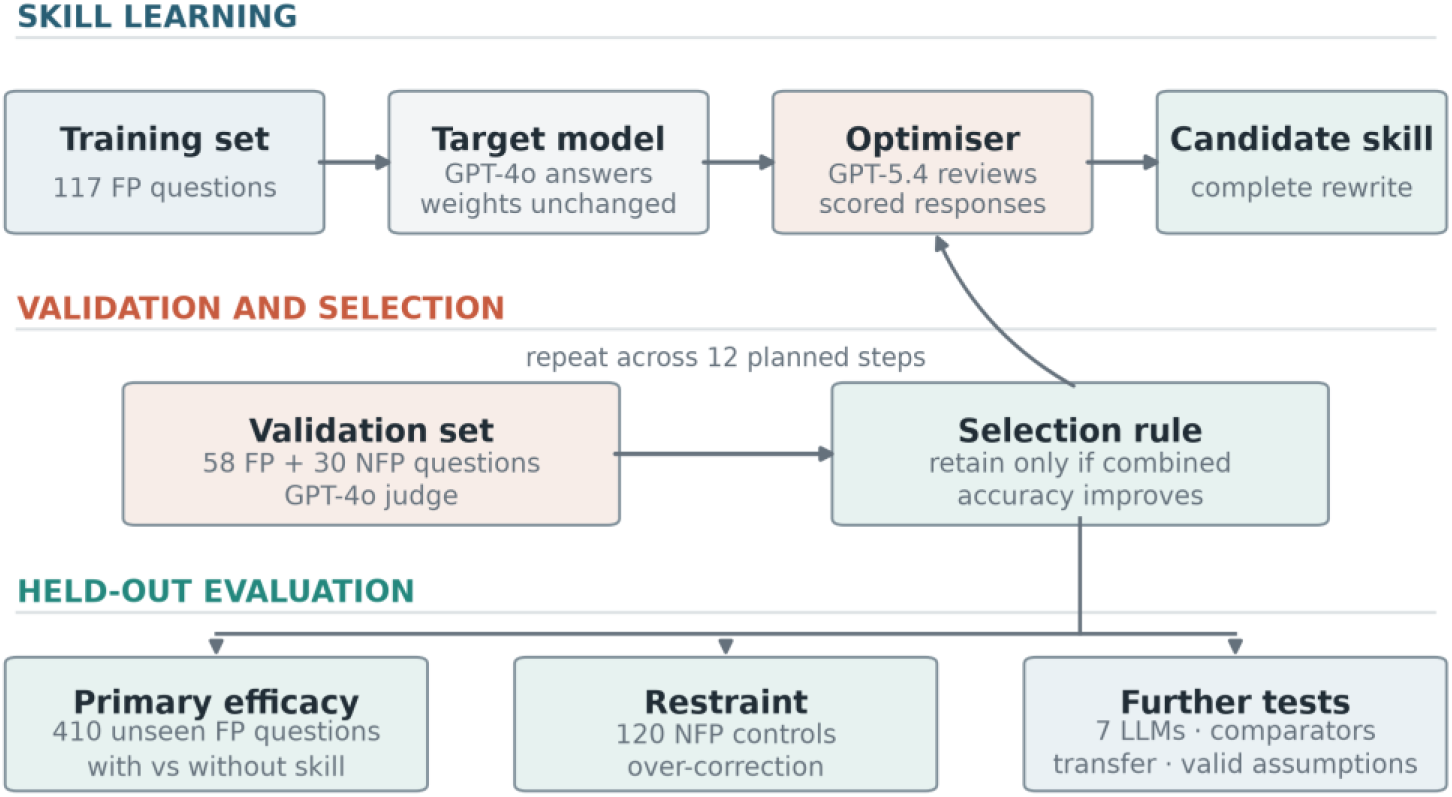
Study design. SkillOpt used GPT-5.4 to review scored GPT-4o responses and rewrite a written skill. A separate GPT-4o judge scored a validation set of 58 FP and 30 NFP questions; a rewrite was retained only when combined validation accuracy improved. The selected skill was tested on 410 unseen FP questions. A standardised evaluation then assessed NFP accuracy, conventional prompts and portability across seven LLMs.

**Table 1.**
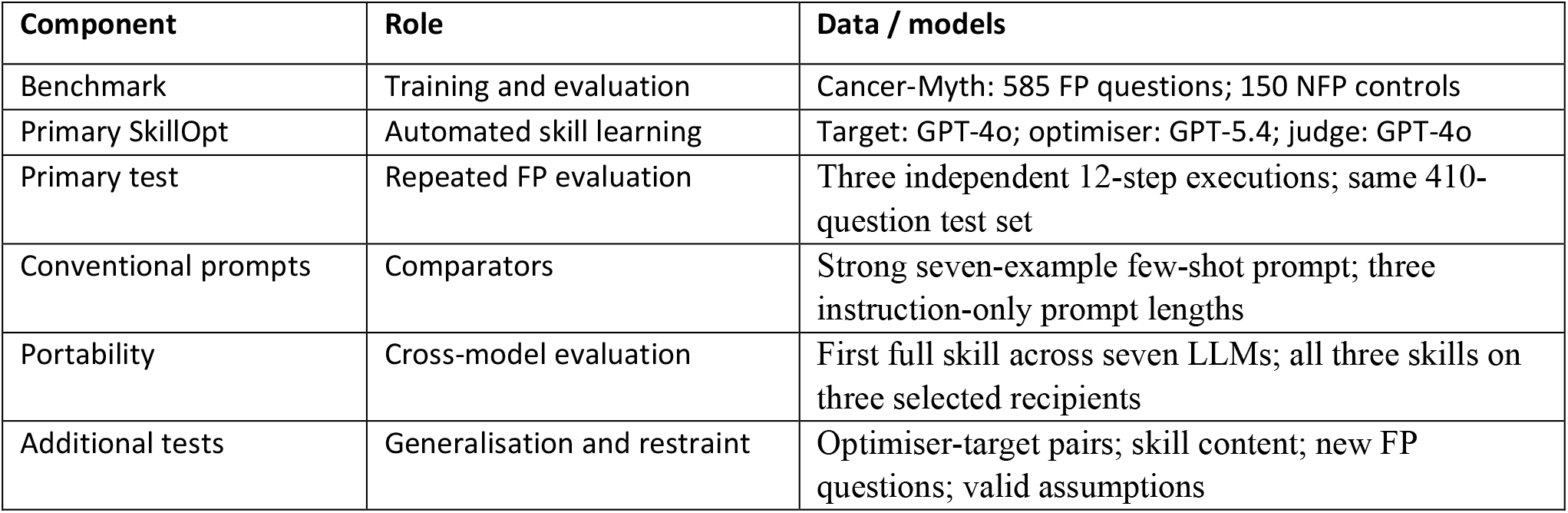
Study components and evaluation populations. The primary analysis comprised three independent GPT-4o SkillOpt executions. Secondary analyses examined optimiser-target combinations, conventional prompts, skill content, portability, compression, generalisation, valid clinical assumptions and judge sensitivity.

| Component | Role | Data / models |
| --- | --- | --- |
| Benchmark | Training and evaluation | Cancer-Myth: 585 FP questions; 150 NFP controls |
| Primary SkillOpt | Automated skill learning | Target: GPT-4o; optimiser: GPT-5.4; judge: GPT-4o |
| Primary test | Repeated FP evaluation | Three independent 12-step executions; same 410-question test set |
| Conventional prompts | Comparators | Strong seven-example few-shot prompt; three instruction-only prompt lengths |
| Portability | Cross-model evaluation | First full skill across seven LLMs; all three skills on three selected recipients |
| Additional tests | Generalisation and restraint | Optimiser-target pairs; skill content; new FP questions; valid assumptions |

### SkillOpt training and primary evaluation

GPT-4o-2024-11-20 was the target model and GPT-5.4-2026-03-05 was the optimiser. Three independent executions used the same split, seed and 128-character starting instruction; they were repeat executions, not different-seed experiments. Each had 12 planned steps and batches of 40 FP questions. In rewrite-from-suggestions mode, GPT-5.4 received scored success and failure traces and rewrote the complete skill. Full-skill rewrites allowed 2,000 completion tokens, with an initial edit budget of four. A rewrite was retained only if one combined hard-accuracy score improved across all 58 FP and 30 NFP validation questions. The rule rewarded both outcomes but did not impose a separate threshold for either one.

The primary analysis compared GPT-4o with and without the selected skill on the same 410 FP test questions in each execution. The baseline contained no task-specific correction instruction. Native primary records did not contain matched NFP responses, so primary inference addressed FP correction; NFP accuracy was measured in the standardised evaluation below. Execution 1 was continued to 24 steps as an exploratory convergence check, but the main learning analysis reports the 12 planned steps. The continuation did not change its selected skill and is detailed in the appendix.

### Cross-model comparisons

A standardised harness evaluated seven LLMs: Llama 3.1 8B Instruct, Gemini 2.5 Flash, Llama 3.3 70B Instruct, GPT-4o, GPT-5.4, Gemini 3.5 Flash and GPT-5.6 Luna. Cross-model analyses used the same 410 FP test questions and 120 NFP controls. Missing responses and invalid judge scores were counted as failures; available-case estimates are reported in the appendix.

We compared three reproducible prompt strategies with no task-specific correction instruction. First, GPT-5.4 generated a deliberately strong 5,927-character few-shot prompt once at temperature 0. It selected five FP examples from 30 candidates in the training split and two NFP examples from 20 candidate controls, then wrote exemplar responses. It was blinded to SkillOpt and the prompt was approximately size-matched to a full skill. Second, GPT-5.4 generated instruction-only prompts once at temperature 0 using 30 FP training examples as task context. The same template produced an unconstrained prompt and prompts targeting approximately 5,000 and 2,000 characters; final lengths were 14,533, 5,774 and 2,814 characters. The prompts contained rules but no worked examples. Third, we evaluated the first full GPT-4o SkillOpt skill and a 1,969-character checkpoint from a separate length-constrained run with a 1,000-token rewrite limit. We excluded bespoke human-written prompts because author wording would add an uncontrolled comparator that could not be standardised or reproduced.

### Secondary analyses

We prespecified eight features for comparison across the three primary skills: premise checking; correction before advice; an explicit instruction to state that an incorrect assumption was wrong and replace it with accurate framing; conditional language; clinical determinants; oncology-team referral; avoidance of guidance that reinforced the FP; and supportive tone. A feature counted as present only when explicitly stated.

Secondary 12-step experiments varied the optimiser and target: GPT-5.6 Luna optimising GPT-4o, GPT-5.4 self-optimisation, GPT-4o self-optimisation and GPT-5.4 optimising Gemini 2.5 Flash. Incomplete runs are listed in the appendix and were not interpreted as failed optimisation. For principal portability testing, the first GPT-4o skill was applied unchanged to the other six models. All three independently learned GPT-4o skills were additionally compared on three selected recipients only: Llama 3.3 70B, GPT-5.4 and Gemini 3.5 Flash.

Generalisation used three human-verified sets: 50 new FP questions in the Cancer-Myth format; 25 direct and 25 non-cancer FP questions; and 25 questions built on valid clinical assumptions. GPT-4o, GPT-5.4 and Llama 3.3 70B were tested with no correction instruction, few-shot prompting and the first GPT-4o skill. A GPT-4o judge applied the Cancer-Myth rubric: FP correction was the proportion scored +1 on a -1, 0, +1 scale; NFP responses received +1 when no FP was invented and -1 otherwise. A stratified 45-response sample and a separate 50-response high-scoring sample were rescored by alternative judges. Existing human-review records were excluded pending reconciliation of their sampling and labels.

### Statistical analysis

Proportions are reported with two-sided 95% Wilson confidence intervals. Within each primary execution, absolute differences used outcomes on the same 410 questions; 95% confidence intervals were estimated from 10,000 paired bootstrap samples. Exact two-sided McNemar tests compared discordant pairs and were adjusted across the three primary comparisons using Holm’s method. Other analyses were secondary or exploratory.

## Results

### Repeated SkillOpt training on GPT-4o

Without a learned skill, GPT-4o corrected 16.3%, 17.6% and 15.4% of FP questions across the three independent executions. With SkillOpt, correction increased to 82.7%, 90.5% and 82.2%, respectively (Table 2; Figure 2A). Absolute improvements were 66.3, 72.9 and 66.8 percentage points; their 95% bootstrap confidence intervals ranged from 61.7 to 77.3 percentage points, and all Holm-adjusted McNemar p values were <0.001.

**Table 2.**
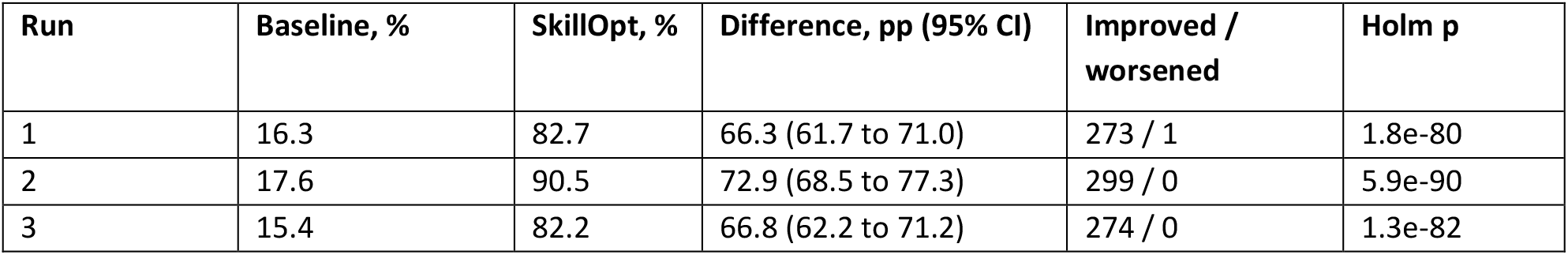
GPT-4o performance with and without the SkillOpt skill across three independent executions. Results use the same 410 FP test questions in each comparison. Condition-specific confidence intervals are Wilson intervals; intervals for absolute differences use 10,000 paired bootstrap samples.

**Figure 2.**
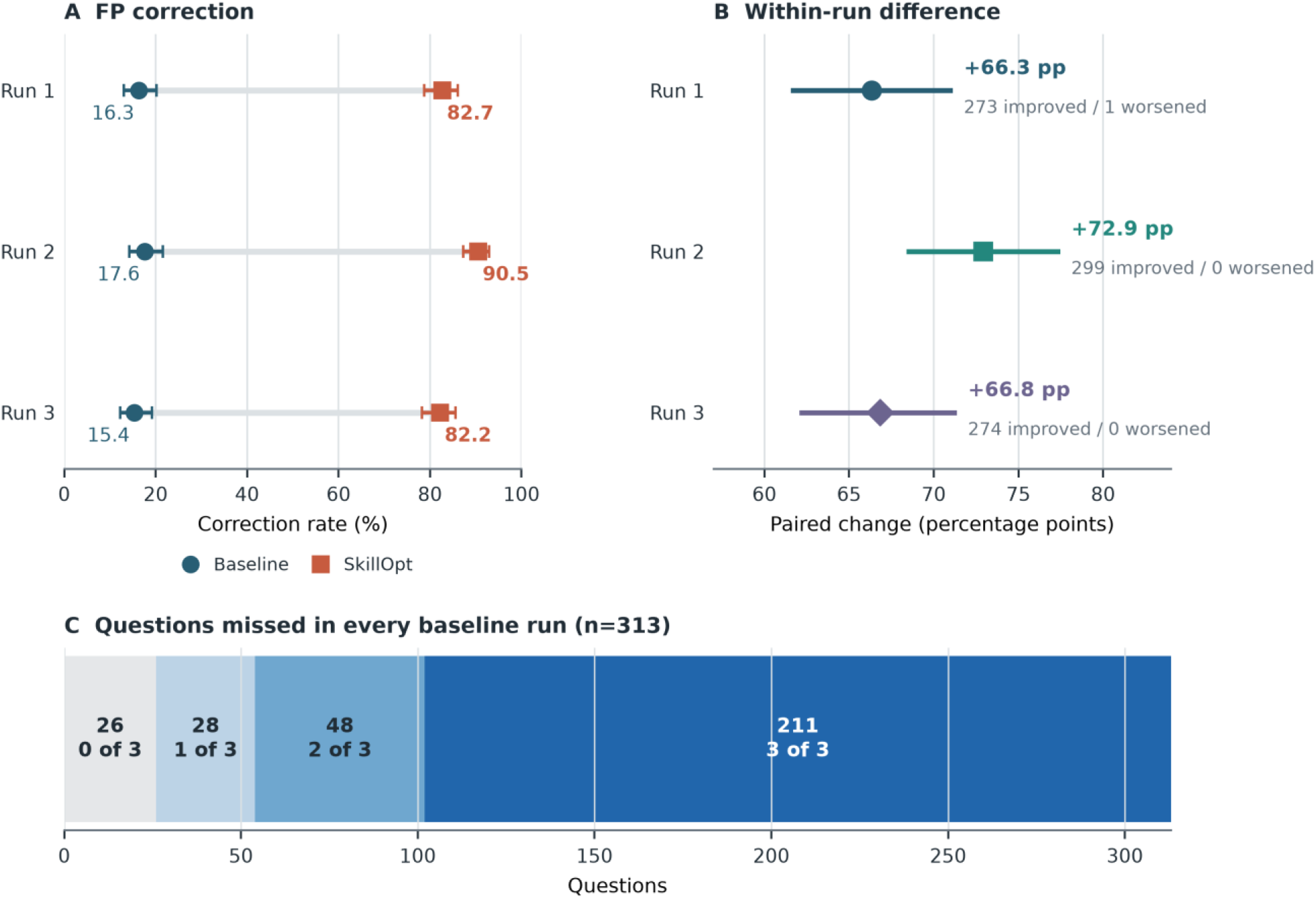
Repeated improvement in FP correction after SkillOpt training. Panel A compares correction with and without the learned skill in each independent execution. Panel B shows absolute within-question differences and 95% bootstrap confidence intervals. Panel C summarises outcomes for the 313 questions missed in every baseline execution.

The changes were strongly directional. Across the three executions, 273, 299 and 274 questions changed from failure to success, whereas one, zero and zero changed in the opposite direction (Figure 2B). Of 313 questions missed in every baseline execution, 211 were corrected in all three SkillOpt executions, 48 in two, 28 in one and 26 in none (Figure 2C). Correction improved in all 21 execution-by-category analyses, with gains of 46.2–83.3 percentage points.

Only two, three and two rewrites were accepted across the primary executions. The best skill appeared by step 3 in executions 1 and 3 and at step 12 in execution 2 (Figure 3A). The exploratory continuation of execution 1 did not improve its step-3 skill. Secondary training depended on the optimiser-target pair (Figure 3B). GPT-5.6 Luna as optimiser increased GPT-4o correction from 17.6% to 58.8%, while GPT-5.4 self-optimisation increased correction from 59.5% to 89.8%. GPT-4o self-optimisation completed 12 steps without producing a rewrite. GPT-5.4 optimisation of Gemini 2.5 Flash improved correction from 2.0% to 20.5%, but final performance remained low.

**Figure 3.**
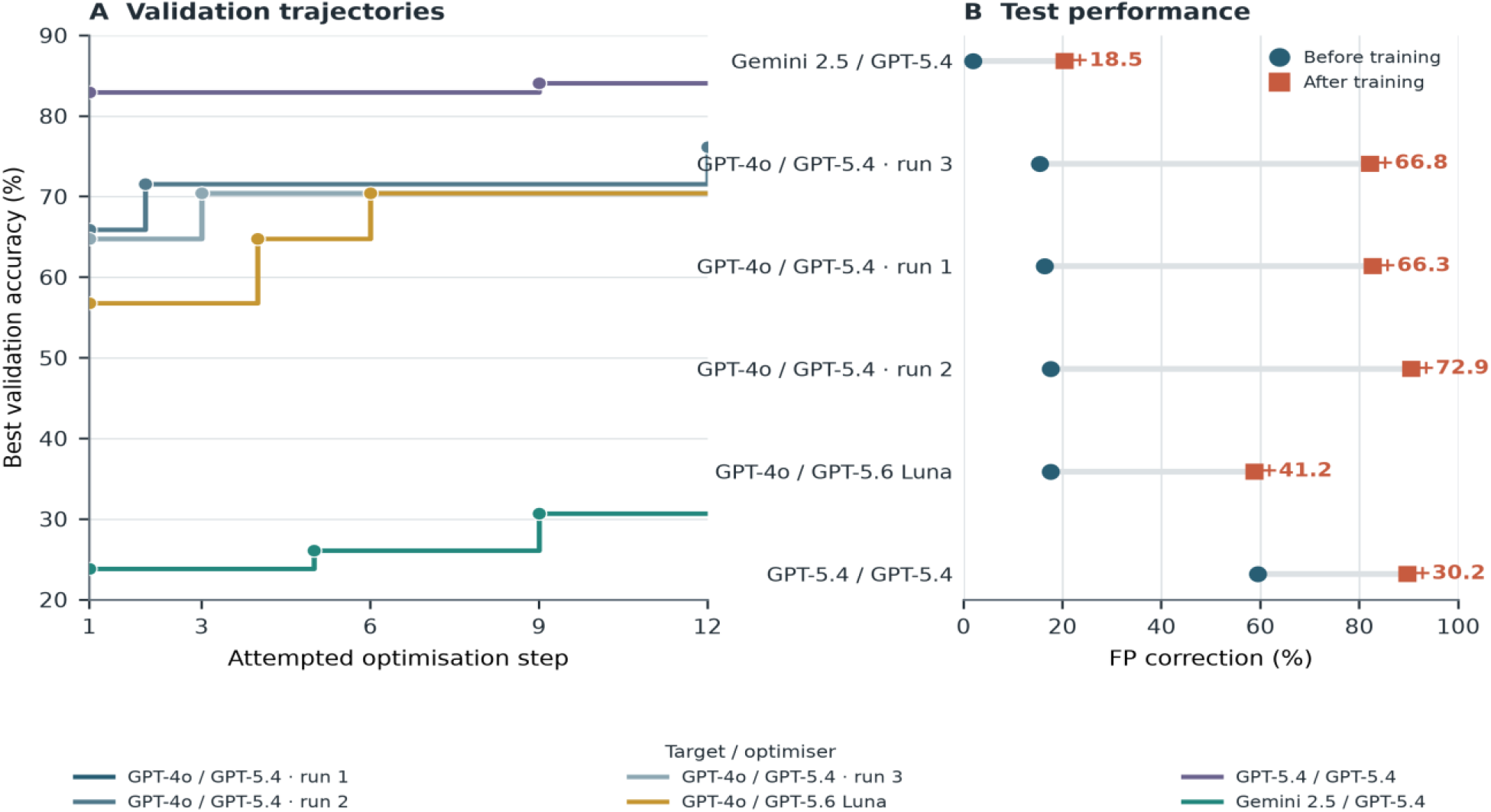
SkillOpt learning dynamics during the 12 planned optimisation steps. Panel A shows the best combined validation accuracy retained after each step; markers indicate accepted rewrites. Panel B compares FP correction before training (blue circles) and after training (orange squares) for completed 12-step experiments. GPT-5.4/GPT-5.4 denotes self-optimisation. The compact run and exploratory post-step-12 continuation are reported in the appendix.

### Skill content and portability

The three primary skills contained 4,933, 4,414 and 5,416 characters. Seven prespecified features appeared in all three: checking the question’s assumption, correcting it before advice, using conditional language, explaining relevant clinical factors, recommending oncology-team input, avoiding advice that reinforced the false assumption, and using supportive language (Figure 4B). Run 2 alone explicitly instructed the model to state that an incorrect assumption was wrong and replace it with accurate framing. Independent executions therefore learned a common communication sequence, with one potentially important difference.

**Figure 4.**
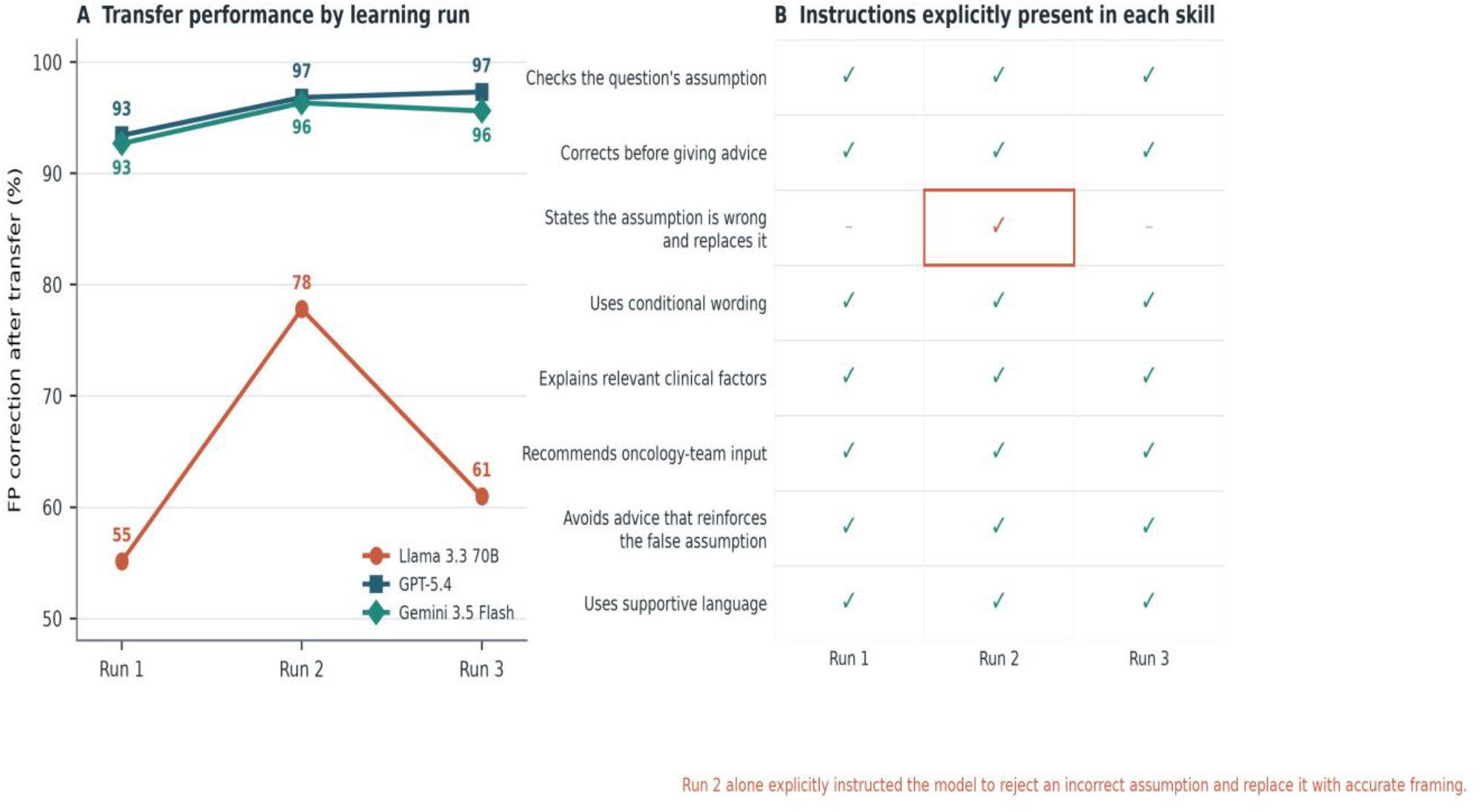
Transfer performance and content of independently learned skills. Panel A compares FP correction after transferring all three primary GPT-4o skills to Llama 3.3 70B, GPT-5.4 and Gemini 3.5 Flash. Panel B shows whether each plain-language instruction was explicitly present (tick) or absent (dash). Run 2, the first replicate, was the only skill that explicitly instructed the model to reject an incorrect assumption and replace it with accurate framing; it also produced the highest Llama 3.3 70B correction. This association does not establish causation.

The first full GPT-4o skill was evaluated more broadly. On its target GPT-4o, FP correction was 82.7% in primary execution 1. A separate standardised-harness evaluation of the same skill recorded 70.0% NFP accuracy on GPT-4o. After unchanged transfer, correction reached 93.4% on GPT-5.4, 92.7% on Gemini 3.5 Flash and 96.6% on GPT-5.6 Luna. It was lower on Llama 3.3 70B (55.1%) and poor on Llama 3.1 8B (16.3%) and Gemini 2.5 Flash (6.3%) (Table 3; Figure 5A). NFP accuracy was 84.2% on GPT-5.4, 79.2% on GPT-5.6 Luna, 59.2% on Gemini 3.5 Flash and 50.0% on Llama 3.3 70B.

**Table 3.**
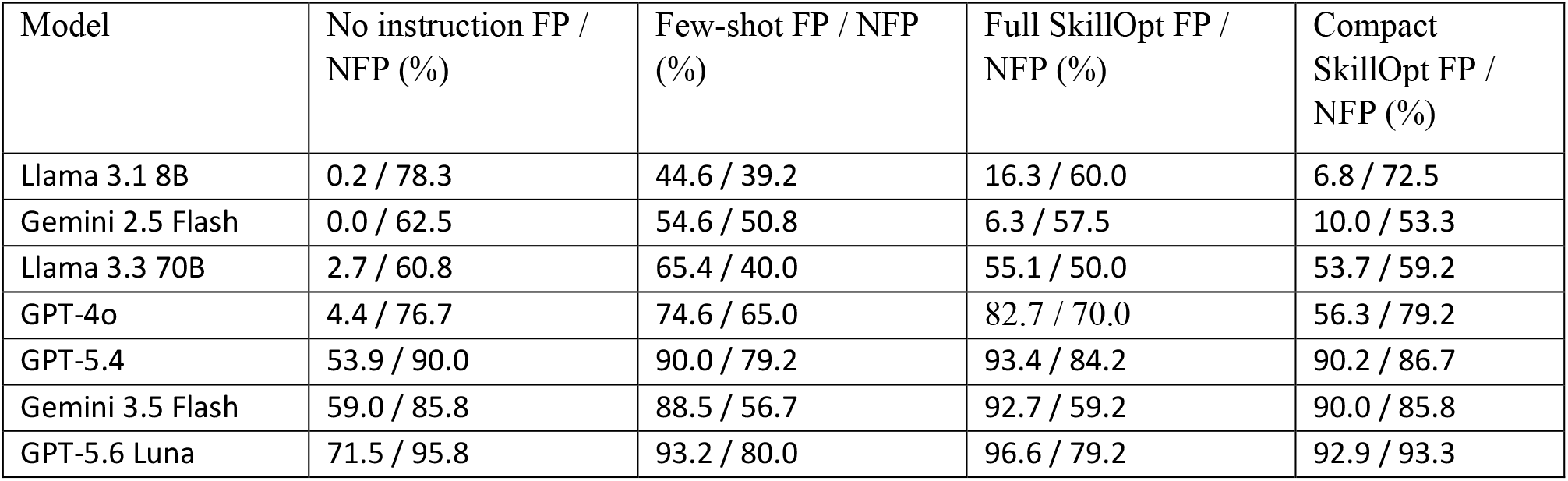
FP correction and NFP accuracy across models and prompt conditions. All values are percentages on the common 410-question FP test set and 120 reserved NFP controls. For GPT-4o full SkillOpt, FP correction (82.7%) is from primary execution 1 and NFP accuracy (70.0%) is from the standardised-harness evaluation of the same skill because primary NFP records were not available. Missing or invalid responses count as failures. Confidence intervals and available-case estimates are reported in the appendix.

| Model | No instruction FP / NFP (%) | Few-shot FP / NFP (%) | Full SkillOpt FP / NFP (%) | Compact SkillOpt FP / NFP (%) |
| --- | --- | --- | --- | --- |
| Llama 3.1 8B | 0.2 / 78.3 | 44.6 / 39.2 | 16.3 / 60.0 | 6.8 / 72.5 |
| Gemini 2.5 Flash | 0.0 / 62.5 | 54.6 / 50.8 | 6.3 / 57.5 | 10.0 / 53.3 |
| Llama 3.3 70B | 2.7 / 60.8 | 65.4 / 40.0 | 55.1 / 50.0 | 53.7 / 59.2 |
| GPT-4o | 4.4 / 76.7 | 74.6 / 65.0 | 82.7 / 70.0 | 56.3 / 79.2 |
| GPT-5.4 | 53.9 / 90.0 | 90.0 / 79.2 | 93.4 / 84.2 | 90.2 / 86.7 |
| Gemini 3.5 Flash | 59.0 / 85.8 | 88.5 / 56.7 | 92.7 / 59.2 | 90.0 / 85.8 |
| GPT-5.6 Luna | 71.5 / 95.8 | 93.2 / 80.0 | 96.6 / 79.2 | 92.9 / 93.3 |

**Figure 5.**
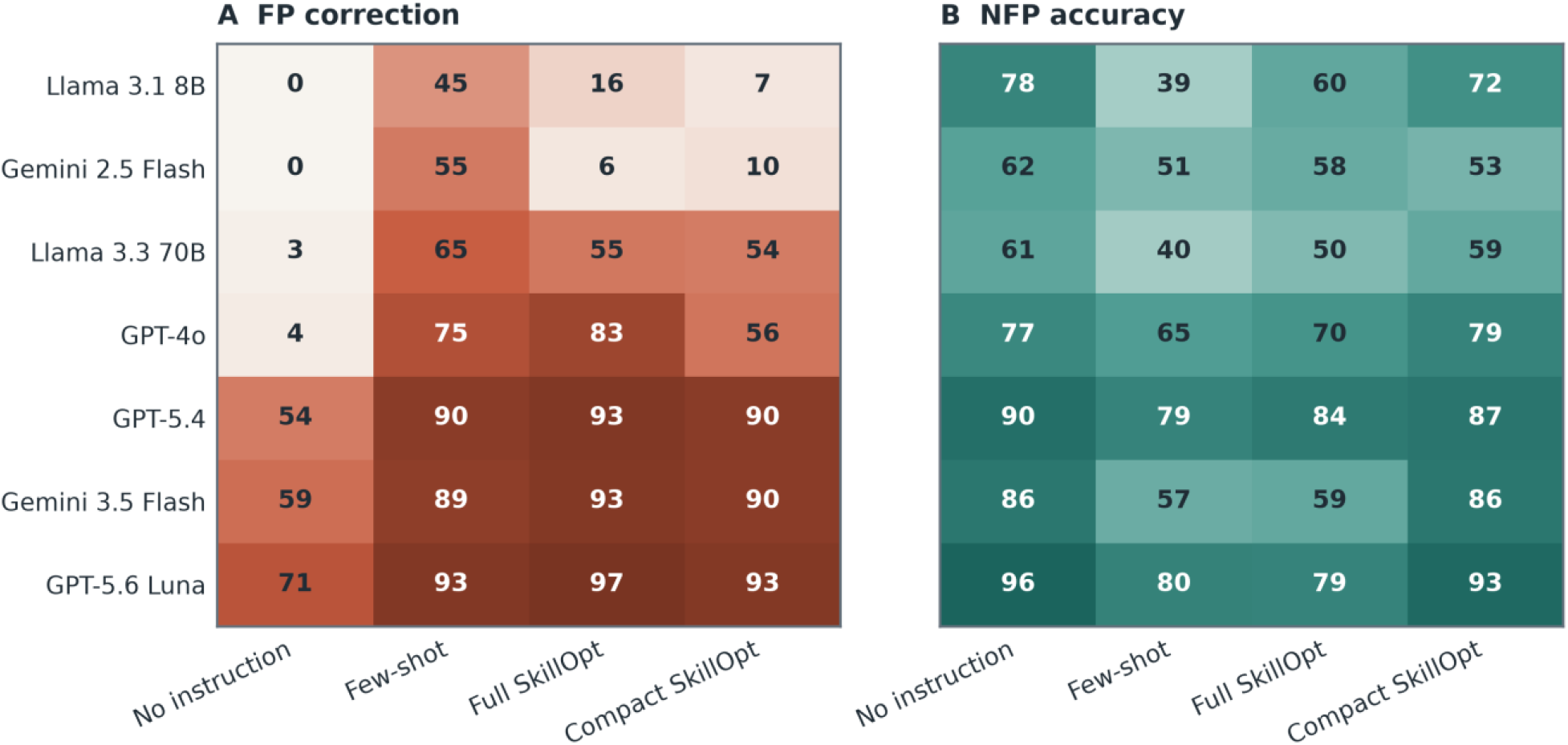
FP correction and NFP accuracy across seven LLMs. Heatmaps compare no correction instruction, the deliberately strong seven-example few-shot prompt, the first full GPT-4o SkillOpt skill and compact SkillOpt. Values are percentages. For GPT-4o full SkillOpt, the FP value is from primary execution 1 and the NFP value is from the standardised-harness evaluation of the same skill.

All three primary skills were transferred only to Llama 3.3 70B, GPT-5.4 and Gemini 3.5 Flash. Performance was similar on GPT-5.4 (93.4–97.3%) and Gemini 3.5 Flash (92.7–96.3%) but diverged on Llama 3.3 70B (55.1%, 77.8% and 61.0%; Figure 4A). The highest result came from Run 2, the only skill that explicitly instructed the model to reject an incorrect assumption and replace it with accurate framing. This difference may explain the stronger Llama 3.3 70B transfer, but the association is hypothesis-generating and does not establish causation.

### Comparison with conventional prompt engineering

Prompt form and length interacted with the receiving model (Figure 6). On Llama 3.1 8B, FP correction was 9.0% with the 14,533-character instruction-only prompt, 15.4% with the 5,774-character prompt, 40.7% with the 2,814-character prompt and 44.6% with few-shot prompting. Llama 3.3 70B showed the same ordering: 26.1%, 40.5%, 42.2% and 65.4%. More capable models were less sensitive to prompt length: GPT-5.4 and Gemini 3.5 Flash achieved 85.6– 92.4% correction with all three instruction-only prompts.

**Figure 6.**
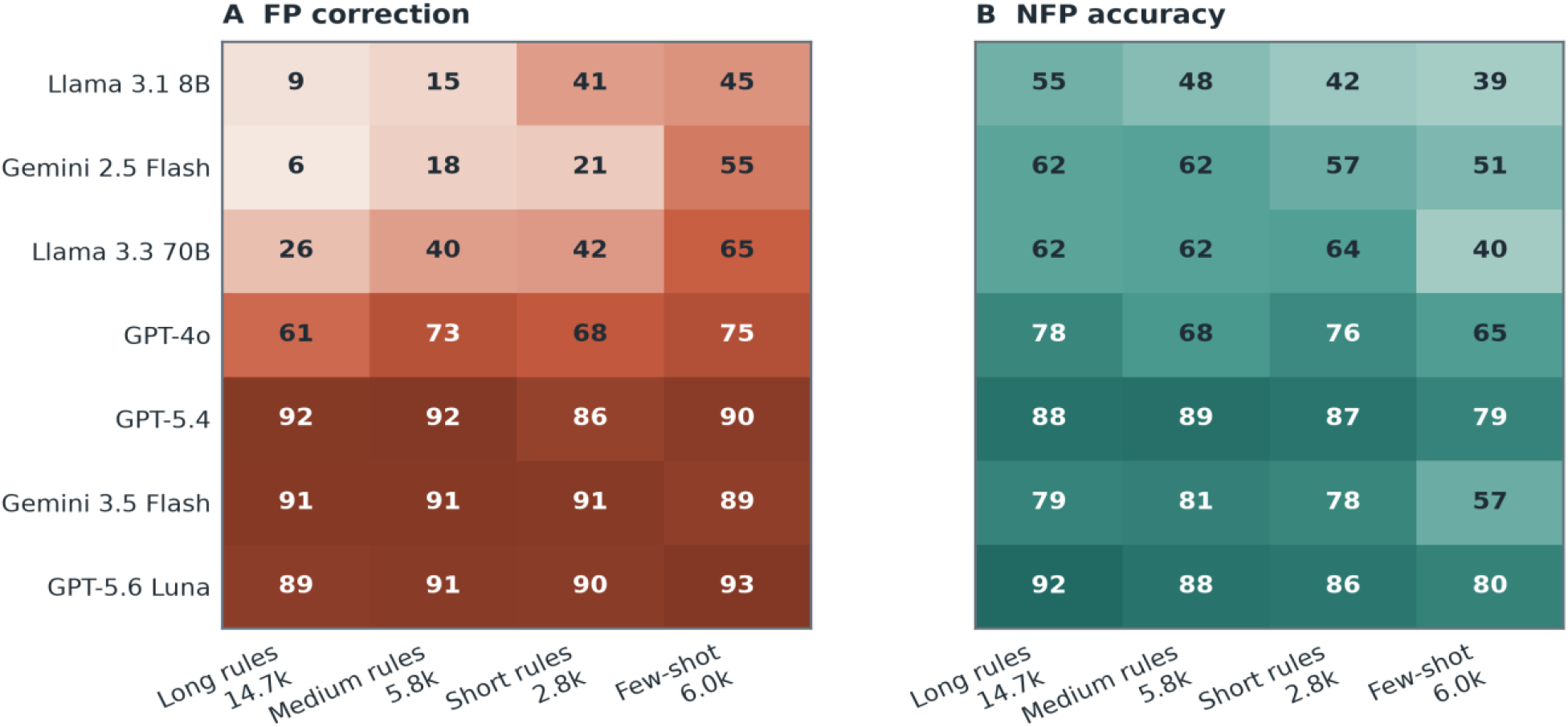
Effect of prompt form and length across LLMs. Three GPT-5.4-generated instruction-only prompts containing no worked examples are compared with the seven-example few-shot prompt. Separate panels show FP correction and NFP accuracy as percentages.

Higher FP correction often reduced NFP accuracy. On Llama 3.1 8B, few-shot prompting increased correction from 0.2% to 44.6%, while NFP accuracy fell from 78.3% to 39.2%. On Gemini 3.5 Flash, few-shot prompting and full SkillOpt achieved 88.5% and 92.7% correction, but NFP accuracy fell from 85.8% with no instruction to 56.7% and 59.2%. Combining the full skill with worked examples was more aggressive: on GPT-4o it achieved 87.6% FP correction but 48.3% NFP accuracy; on Gemini 2.5 Flash the corresponding values were 48.3% and 45.0%.

Compact SkillOpt improved this balance on several capable models. On Gemini 3.5 Flash, it retained 90.0% FP correction and increased NFP accuracy from 59.2% with the full skill to 85.8%. On GPT-5.6 Luna, correction was 92.9% and NFP accuracy increased from 79.2% to 93.3%. GPT-5.4 showed a smaller change: correction fell from 93.4% to 90.2% and NFP accuracy increased from 84.2% to 86.7%. Compression was not universally beneficial; Llama 3.1 8B achieved only 6.8% FP correction with compact SkillOpt.

### Generalisation and valid clinical assumptions

On 50 new FP questions in the Cancer-Myth format, the first SkillOpt skill improved correction from 2% to 80% on GPT-4o, from 42% to 90% on GPT-5.4 and from 6% to 62% on Llama 3.3 70B (Figure 7A). Few-shot results were 76%, 92% and 66%. On 50 differently framed FP questions, including non-cancer topics, SkillOpt achieved 98–100% correction across the three models (Figure 7B). High baseline performance for GPT-4o and GPT-5.4 on this set limited separation between interventions.

**Figure 7.**
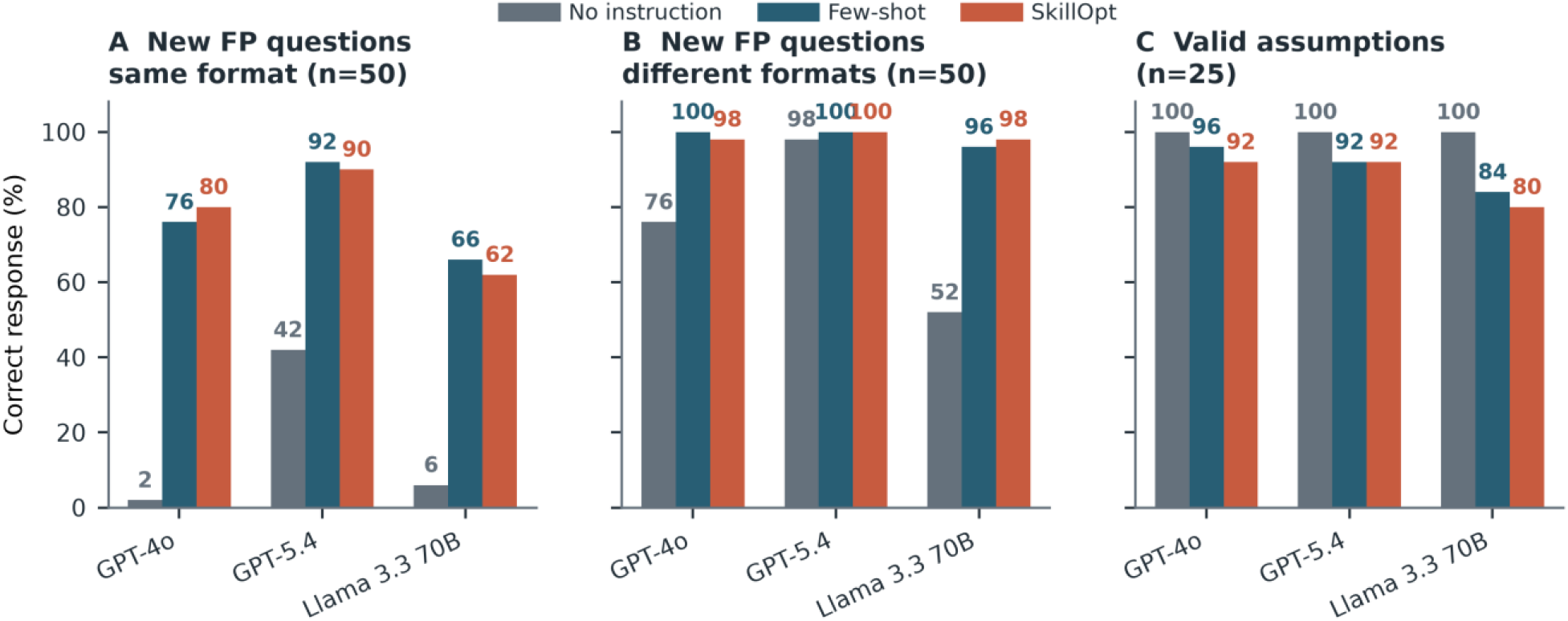
Performance on new FP questions and questions with valid clinical assumptions. Panels A and B show correction on two human-verified sets of 50 new FP questions. Panel C shows the percentage of 25 human-verified valid-assumption questions answered without inappropriate correction. Values above bars are percentages.

On 25 questions built on valid clinical assumptions, every baseline response avoided inappropriate correction. With SkillOpt, appropriate responses fell to 92% on GPT-4o and GPT-5.4 and 80% on Llama 3.3 70B. Few-shot prompting produced 96%, 92% and 84%, respectively (Figure 7C). Both interventions therefore retained a measurable risk of over-correction.

Alternative automated judges preserved the direction of the main effects but differed on borderline responses. On a stratified 45-response sample, exact agreement with GPT-4o ranged from 71.1% to 84.4%, and FP correction estimates differed by -6.7 to +8.8 percentage points. On a separate 50-response high-scoring sample, GPT-5.4 and GPT-5.6 Luna agreed with GPT-4o on 96% and 98% of scores, respectively (Appendix Table S17).

## Discussion

SkillOpt produced large, repeatable improvements in GPT-4o’s handling of false presuppositions. Three independent executions increased correction by 66.3–72.9 percentage points on the same unseen questions, with only two or three accepted rewrites in each. The same difficult questions often improved across executions, and independently learned skills converged on a common sequence: identify the false assumption, correct it before advice, explain the relevant clinical factors and avoid guidance that reinforces the premise.

These findings extend SkillOpt beyond the agentic and reasoning tasks for which it was developed. The target behaviour was not execution of a fixed procedure; it required deciding whether correction was warranted. Cancer-Myth had already shown that Genetic-Pareto optimisation could raise FP correction while causing frequent inappropriate correction of NFP questions [6]. SkillOpt included both outcomes during validation and produced large primary gains, but the native primary records did not include NFP controls, so those gains alone do not show that over-correction was solved.

Learning and transfer depended on the models involved. GPT-5.4 improved itself and produced the strongest GPT-4o skills, whereas GPT-4o generated no rewrite during self-optimisation and GPT-5.6 Luna was a weaker optimiser for GPT-4o. The first GPT-4o skill transferred effectively to GPT-5.4, Gemini 3.5 Flash and GPT-5.6 Luna but poorly to Gemini 2.5 Flash and Llama 3.1 8B. The written artifact was portable across providers, but not model agnostic; a recipient model still had to interpret and apply it.

The conventional comparators distinguish iterative learning from prompt length alone. The few-shot prompt was intentionally strong, using benchmark-derived FP and NFP examples with responses generated by GPT-5.4. The instruction-only prompts were also generated reproducibly rather than written by the investigators. On the Llama models, worked examples and shorter rules outperformed long instructions; more capable models were less sensitive to length. One elaborate prompt is therefore not an adequate universal control condition.

Skill compression changed the balance between correction and restraint. On Gemini 3.5 Flash and GPT-5.6 Luna, compact SkillOpt preserved most FP correction while substantially improving NFP accuracy. Longer skills may contain instructions that capable models over-apply, although these experiments cannot identify which removed instructions caused the change. Compression did not overcome weak instruction following on the least responsive models and remains a model-specific intervention.

The valid-assumption tests expose the practical limit of both automated skills and few-shot prompting. Each sometimes contradicted a premise that should have been accepted, with the largest effect on Llama 3.3 70B. A prompt should not be described as a safety layer because it improves FP correction alone. For clinical AI development, the useful unit is the model-prompt combination: model version, prompt version, FP test set, NFP controls and valid-assumption tests should be recorded and evaluated together.

Strengths include within-question primary comparisons repeated across three independent executions; common FP and NFP populations for cross-model analyses; conservative handling of missing outputs; and retained prompt artifacts and source hashes. The programme also tested conventional prompts, multiple optimiser-target pairs, independently learned skills, cross-provider transfer, compression, new questions and alternative judges.

The primary executions used the same split and seed, so they establish repeatability rather than robustness to alternative partitions. Most secondary conditions generated one response per question and do not estimate repeated sampling variation. The combined validation score did not impose separate FP and NFP thresholds. GPT-4o was the main judge, and alternative judges disagreed on some borderline responses. Existing human-review records were excluded because their sampling and labels require reconciliation. Outcomes measured FP correction and NFP accuracy, not overall factual accuracy, empathy, comprehension or clinical harm. Proprietary model behaviour may also change after provider updates.

In conclusion, SkillOpt can discover inspectable instructions that substantially improve a difficult clinical communication behaviour. Its value here was not a universal prompt, but a reproducible process for learning and testing model-specific behaviour. SkillOpt outperformed conventional prompts on capable models, few-shot prompting remained stronger on some less responsive models, and over-correction persisted. Prompts should therefore be selected and versioned for each model using paired measures of correction and restraint.

## Data Availability

All data produced in the present study are available upon reasonable request to the authors

## Declarations

### Ethics

This study analysed an existing benchmark and model-generated responses. It involved no research participants or identifiable personal data; formal research ethics approval was therefore not required.

### Data and code availability

Cancer-Myth is available under the terms set by its authors. The analysis package contains prompt and skill artifacts, derived tables, source-file hashes and reproducible scripts. Release of complete model responses remains subject to benchmark licensing, model-provider terms and institutional review.

