## Supplemental Data for "Automated Skill Optimisation for False Presupposition Handling in Cancer Communication: SkillOpt Versus Conventional Prompt Engineering Across Language Models"

Sanjay Khanna, MBBS MRCP

#### S1. Study reconstruction and source integrity

The publication analysis was reconstructed from the original experiment directories without changing source files. A recursive audit identified 153 JSONL response ledgers and three record schemas. Each included ledger was linked to its path and SHA-256 hash. One malformed legacy Llama 3.1 8B baseline contained duplicate records and was excluded in favour of the later canonical 585-record ledger. Twenty-seven otherwise usable ledgers contained missing or invalid scores; these were retained using the prespecified missing-as-failure analysis, with available-case estimates reported alongside it.

The primary analysis used six complete GPT-4o ledgers: baseline and SkillOpt responses from each of three runs. Each ledger contained the same 410 question IDs, and question text was identical within every comparison. Record-derived scores reproduced the saved summaries.

##### Table S1. Complete evaluation-ledger registry

| Family | Condition | Type | Model | Rows / scored | Status | Finding | SHA-256 |
|---|---|---|---|---|---|---|---|
| Baseline capability | baseline_5_4_gpt-5.4_fp_run1 | fp | gpt-5.4 | 585 / 585 | usable | no record-level integrity findings | 1607587ca886… |
| Baseline capability | baseline_5_4_gpt-5.4_nfp_run1 | nfp | gpt-5.4 | 150 / 150 | usable | no record-level integrity findings | 327cb2d2e13e… |
| Baseline capability | baseline_gemini-2.5-flash_fp_run1 | fp | gemini-2.5-flash | 585 / 573 | usable with conservative missing-as-failure analysis | invalid_scores=12 | 0262b9ba742f… |
| Baseline capability | baseline_gemini-2.5-flash_nfp_run1 | nfp | gemini-2.5-flash | 150 / 146 | usable with conservative missing-as-failure analysis | invalid_scores=4 | b40f13ae62ad… |
| Baseline capability | baseline_gemini-3.5-flash_fp_run1 | fp | gemini-3.5-flash | 585 / 585 | usable | no record-level integrity findings | 66eb3a55b43d… |
| Baseline capability | baseline_gemini-3.5-flash_nfp_run1 | nfp | gemini-3.5-flash | 150 / 150 | usable | no record-level integrity findings | cf897f61b579… |
| Baseline capability | baseline_gpt-4o-2024-11-20_fp_run1 | fp | gpt-4o-2024-11-20 | 585 / 585 | usable | no record-level integrity findings | e2c970dea97d… |
| Baseline capability | baseline_gpt-4o-2024-11-20_nfp_run1 | nfp | gpt-4o-2024-11-20 | 150 / 150 | usable | no record-level integrity findings | 395fcd2d1612… |
| Baseline capability | baseline_llama-3.1-8b_fp_run1 | fp | llama-3.1-8b | 693 / 583 | exclude pending repair | duplicate_qids=108; target_errors=110; judge_errors=110; blank_responses=110; invalid_scores=110; nonstandard_rows=693 | ce9ce116fd1b… |
| Baseline capability | baseline_llama-3.1-8b_nfp_run1 | nfp | llama-3.1-8b | 150 / 150 | usable | no record-level integrity findings | 157533b0702e… |
| Baseline capability | baseline_luna_fp_run1 | | | 54 / 54 | usable with conservative missing-as-failure analysis | nonstandard_rows=54 | 00ceae580522… |
| Baseline capability | baseline_luna_gpt-5.6-luna_fp_run1 | fp | gpt-5.6-luna | 585 / 585 | usable | no record-level integrity findings | bd3ed1f8a3c8… |
| Baseline capability | baseline_luna_gpt-5.6-luna_nfp_run1 | nfp | gpt-5.6-luna | 150 / 150 | usable | no record-level integrity findings | da8e906d8d5b… |
| Baseline capability | baseline_luna_nfp_run1 | | | 3 / 3 | usable with conservative missing-as-failure analysis | nonstandard_rows=3 | 9802cc3484db… |
| Baseline capability | baseline_us.meta.llama3-1-8b-instruct-v1:0_fp_run1 | fp | us.meta.llama3-1-8b-instruct-v1:0 | 585 / 585 | usable | no record-level integrity findings | b536db4b269a… |
| Baseline capability | baseline_us.meta.llama3-1-8b-instruct-v1:0_nfp_run1 | nfp | us.meta.llama3-1-8b-instruct-v1:0 | 150 / 150 | usable | no record-level integrity findings | 538481e2bfc0… |
| Baseline capability | baseline_us.meta.llama3-3-70b-instruct-v1:0_fp_run1 | fp | us.meta.llama3-3-70b-instruct-v1:0 | 585 / 585 | usable | no record-level integrity findings | ad6f74e0907a… |
| Baseline capability | baseline_us.meta.llama3-3-70b-instruct-v1:0_nfp_run1 | nfp | us.meta.llama3-3-70b-instruct-v1:0 | 150 / 150 | usable | no record-level integrity findings | ce4664da2fd1… |
| Composite intervention | comp_gemini_gemini-2.5-flash_fp_run1 | fp | gemini-2.5-flash | 585 / 569 | usable with conservative missing-as-failure analysis | invalid_scores=16 | 9647edb05fe3… |
| Composite intervention | comp_gemini_gemini-2.5-flash_nfp_run1 | nfp | gemini-2.5-flash | 150 / 150 | usable | no record-level integrity findings | 61272d3e77e0… |
| Composite intervention | comp_gpt4o_gpt-4o-2024-11-20_fp_run1 | fp | gpt-4o-2024-11-20 | 585 / 585 | usable | no record-level integrity findings | 9f30cadb064e… |
| Composite intervention | comp_gpt4o_gpt-4o-2024-11-20_nfp_run1 | nfp | gpt-4o-2024-11-20 | 150 / 150 | usable | no record-level integrity findings | df4cb448acbe… |
| Compression | compact_skill_gemini-2.5-flash_fp_run1 | fp | gemini-2.5-flash | 585 / 551 | usable with conservative missing-as-failure analysis | invalid_scores=34 | 43c94262632c… |
| Compression | compact_skill_gemini-2.5-flash_nfp_run1 | nfp | gemini-2.5-flash | 150 / 148 | usable with conservative missing-as-failure analysis | invalid_scores=2 | cfc8815a2e75… |
| Compression | compact_skill_gemini-3.5-flash_fp_run1 | fp | gemini-3.5-flash | 585 / 585 | usable | no record-level integrity findings | f7718ee1e19d… |
| Compression | compact_skill_gemini-3.5-flash_nfp_run1 | nfp | gemini-3.5-flash | 150 / 150 | usable | no record-level integrity findings | 9374ef5d8111… |
| Compression | compact_skill_gpt-4o-2024-11-20_fp_run1 | fp | gpt-4o-2024-11-20 | 585 / 585 | usable | no record-level integrity findings | caabed18f0ef… |

| Family | Condition | Type | Model | Rows / scored | Status | Finding | SHA-256 |
|---|---|---|---|---|---|---|---|
| Compression | compact_skill_gpt-4o-2024-11-20_nfp_run1 | nfp | gpt-4o-2024-11-20 | 150 / 150 | usable | no record-level integrity findings | d5f0eb03536e… |
| Compression | compact_skill_gpt-5.4_fp_run1 | fp | gpt-5.4 | 585 / 585 | usable | no record-level integrity findings | c42482443049… |
| Compression | compact_skill_gpt-5.4_nfp_run1 | nfp | gpt-5.4 | 150 / 150 | usable | no record-level integrity findings | 6d30787b4251… |
| Compression | compact_skill_gpt-5.6-luna_fp_run1 | fp | gpt-5.6-luna | 585 / 585 | usable | no record-level integrity findings | 8b7d8407c860… |
| Compression | compact_skill_gpt-5.6-luna_nfp_run1 | nfp | gpt-5.6-luna | 150 / 150 | usable | no record-level integrity findings | 120cde872f6c… |
| Compression | compact_skill_us.meta.llama3-1-8b-instruct-v1:0_fp_run1 | fp | us.meta.llama3-1-8b-instruct-v1:0 | 585 / 585 | usable | no record-level integrity findings | 25f32442f893… |
| Compression | compact_skill_us.meta.llama3-1-8b-instruct-v1:0_nfp_run1 | nfp | us.meta.llama3-1-8b-instruct-v1:0 | 150 / 150 | usable | no record-level integrity findings | c6e1009ff4cf… |
| Compression | compact_skill_us.meta.llama3-3-70b-instruct-v1:0_fp_run1 | fp | us.meta.llama3-3-70b-instruct-v1:0 | 585 / 585 | usable | no record-level integrity findings | b662b5080202… |
| Compression | compact_skill_us.meta.llama3-3-70b-instruct-v1:0_nfp_run1 | nfp | us.meta.llama3-3-70b-instruct-v1:0 | 150 / 150 | usable | no record-level integrity findings | f80225b9183a… |
| Generalisation | exp_a_baseline_gpt-4o-2024-11-20_fp_run1 | fp | gpt-4o-2024-11-20 | 50 / 49 | usable with conservative missing-as-failure analysis | target_errors=1; judge_errors=1; blank_responses=1; invalid_scores=1 | f3e23dc2dbb8… |
| Generalisation | exp_a_baseline_gpt-5.4_fp_run1 | fp | gpt-5.4 | 50 / 50 | usable | no record-level integrity findings | 1f2f70707a99… |
| Generalisation | exp_a_baseline_us.meta.llama3-3-70b-instruct-v1:0_fp_run1 | fp | us.meta.llama3-3-70b-instruct-v1:0 | 50 / 50 | usable | no record-level integrity findings | bbbd766aeb3b… |
| Generalisation | exp_a_fewshot_gpt-4o-2024-11-20_fp_run1 | fp | gpt-4o-2024-11-20 | 50 / 49 | usable with conservative missing-as-failure analysis | target_errors=1; judge_errors=1; blank_responses=1; invalid_scores=1 | 79a79de97f78… |
| Generalisation | exp_a_fewshot_gpt-5.4_fp_run1 | fp | gpt-5.4 | 50 / 49 | usable with conservative missing-as-failure analysis | target_errors=1; judge_errors=1; blank_responses=1; invalid_scores=1 | fa5717ce2de4… |
| Generalisation | exp_a_fewshot_us.meta.llama3-3-70b-instruct-v1:0_fp_run1 | fp | us.meta.llama3-3-70b-instruct-v1:0 | 50 / 50 | usable | no record-level integrity findings | 8453198f2c8a… |
| Generalisation | exp_a_skillopt_gpt-4o-2024-11-20_fp_run1 | fp | gpt-4o-2024-11-20 | 50 / 49 | usable with conservative missing-as-failure analysis | target_errors=1; judge_errors=1; blank_responses=1; invalid_scores=1 | d1dc8948d40a… |
| Generalisation | exp_a_skillopt_gpt-5.4_fp_run1 | fp | gpt-5.4 | 50 / 49 | usable with conservative missing-as-failure analysis | target_errors=1; judge_errors=1; blank_responses=1; invalid_scores=1 | c2213048c1b4… |
| Generalisation | exp_a_skillopt_us.meta.llama3-3-70b-instruct-v1:0_fp_run1 | fp | us.meta.llama3-3-70b-instruct-v1:0 | 50 / 50 | usable | no record-level integrity findings | 65b3e40232ff… |
| Generalisation | exp_b2_baseline_gpt-4o-2024-11-20_nfp_run1 | nfp | gpt-4o-2024-11-20 | 25 / 25 | usable | no record-level integrity findings | 8d0c5ba0b730… |
| Generalisation | exp_b2_baseline_gpt-5.4_nfp_run1 | nfp | gpt-5.4 | 25 / 25 | usable | no record-level integrity findings | a412d6a6859d… |
| Generalisation | exp_b2_baseline_us.meta.llama3-3-70b-instruct-v1:0_nfp_run1 | nfp | us.meta.llama3-3-70b-instruct-v1:0 | 25 / 25 | usable | no record-level integrity findings | 8e56cc03e3dd… |
| Generalisation | exp_b2_fewshot_gpt-4o-2024-11-20_nfp_run1 | nfp | gpt-4o-2024-11-20 | 25 / 25 | usable | no record-level integrity findings | cb9c7bf78662… |
| Generalisation | exp_b2_fewshot_gpt-5.4_nfp_run1 | nfp | gpt-5.4 | 25 / 25 | usable | no record-level integrity findings | 3d7b6458e680… |
| Generalisation | exp_b2_fewshot_us.meta.llama3-3-70b-instruct-v1:0_nfp_run1 | nfp | us.meta.llama3-3-70b-instruct-v1:0 | 25 / 25 | usable | no record-level integrity findings | d4e1a5dd67ac… |
| Generalisation | exp_b2_skillopt_gpt-4o-2024-11-20_nfp_run1 | nfp | gpt-4o-2024-11-20 | 25 / 25 | usable | no record-level integrity findings | 9dff6c140d36… |
| Generalisation | exp_b2_skillopt_gpt-5.4_nfp_run1 | nfp | gpt-5.4 | 25 / 25 | usable | no record-level integrity findings | 33b135eeb020… |
| Generalisation | exp_b2_skillopt_us.meta.llama3-3-70b-instruct-v1:0_nfp_run1 | nfp | us.meta.llama3-3-70b-instruct-v1:0 | 25 / 25 | usable | no record-level integrity findings | 33c6893e1bc2… |
| Generalisation | exp_b_baseline_gpt-4o-2024-11-20_fp_run1 | fp | gpt-4o-2024-11-20 | 50 / 50 | usable | no record-level integrity findings | 53bebad64d02… |
| Generalisation | exp_b_baseline_gpt-5.4_fp_run1 | fp | gpt-5.4 | 50 / 50 | usable | no record-level integrity findings | 4b69a6b7d126… |
| Generalisation | exp_b_baseline_us.meta.llama3-3-70b-instruct-v1:0_fp_run1 | fp | us.meta.llama3-3-70b-instruct-v1:0 | 50 / 50 | usable | no record-level integrity findings | 6afe36f083e0… |
| Generalisation | exp_b_fewshot_gpt-4o-2024-11-20_fp_run1 | fp | gpt-4o-2024-11-20 | 50 / 50 | usable | no record-level integrity findings | 99c4a0664025… |
| Generalisation | exp_b_fewshot_gpt-5.4_fp_run1 | fp | gpt-5.4 | 50 / 50 | usable | no record-level integrity findings | 372d09db9a00… |
| Generalisation | exp_b_fewshot_us.meta.llama3-3-70b-instruct-v1:0_fp_run1 | fp | us.meta.llama3-3-70b-instruct-v1:0 | 50 / 50 | usable | no record-level integrity findings | 231e243cdc39… |
| Generalisation | exp_b_skillopt_gpt-4o-2024-11-20_fp_run1 | fp | gpt-4o-2024-11-20 | 50 / 50 | usable | no record-level integrity findings | e197800dcd92… |
| Generalisation | exp_b_skillopt_gpt-5.4_fp_run1 | fp | gpt-5.4 | 50 / 50 | usable | no record-level integrity findings | 6f148af91909… |
| Generalisation | exp_b_skillopt_us.meta.llama3-3-70b-instruct-v1:0_fp_run1 | fp | us.meta.llama3-3-70b-instruct-v1:0 | 50 / 50 | usable | no record-level integrity findings | b1a45fb71f16… |
| Prompt strategy | fewshot_gpt54_gemini-2.5-flash_fp_run1 | fp | gemini-2.5-flash | 585 / 576 | usable with conservative missing-as-failure analysis | invalid_scores=9 | 1c59172f5c30… |
| Prompt strategy | fewshot_gpt54_gemini-2.5-flash_nfp_run1 | nfp | gemini-2.5-flash | 150 / 150 | usable | no record-level integrity findings | 937fa227254c… |
| Prompt strategy | fewshot_gpt54_gemini-3.5-flash_fp_run1 | fp | gemini-3.5-flash | 585 / 585 | usable | no record-level integrity findings | 93a878ce53a8… |

| Family | Condition | Type | Model | Rows / scored | Status | Finding | SHA-256 |
|---|---|---|---|---|---|---|---|
| Prompt strategy | fewshot_gpt54_gemini-3.5-flash_nfp_run1 | nfp | gemini-3.5-flash | 150 / 150 | usable | no record-level integrity findings | 03613e4c53fe… |
| Prompt strategy | fewshot_gpt54_gpt-4o-2024-11-20_fp_run1 | fp | gpt-4o-2024-11-20 | 585 / 585 | usable | no record-level integrity findings | ea9fc5a5e8d7… |
| Prompt strategy | fewshot_gpt54_gpt-4o-2024-11-20_nfp_run1 | nfp | gpt-4o-2024-11-20 | 150 / 150 | usable | no record-level integrity findings | 5d921f4e9498… |
| Prompt strategy | fewshot_gpt54_llama-3.1-8b_fp_run1 | fp | llama-3.1-8b | 585 / 584 | usable with conservative missing-as-failure analysis | judge_errors=1; invalid_scores=1 | 2038292e2c69… |
| Prompt strategy | fewshot_gpt54_llama-3.1-8b_nfp_run1 | nfp | llama-3.1-8b | 150 / 150 | usable | no record-level integrity findings | c70fcbdef35f… |
| Prompt strategy | fewshot_gpt54_us.meta.llama3-1-8b-instruct-v1:0_fp_run1 | fp | us.meta.llama3-1-8b-instruct-v1:0 | 585 / 585 | usable | no record-level integrity findings | 3d3cb44af552… |
| Prompt strategy | fewshot_gpt54_us.meta.llama3-1-8b-instruct-v1:0_nfp_run1 | nfp | us.meta.llama3-1-8b-instruct-v1:0 | 150 / 150 | usable | no record-level integrity findings | 9e083dde0987… |
| Prompt strategy | fewshot_gpt54_us.meta.llama3-3-70b-instruct-v1:0_fp_run1 | fp | us.meta.llama3-3-70b-instruct-v1:0 | 585 / 585 | usable | no record-level integrity findings | 4e0f52ca326c… |
| Prompt strategy | fewshot_gpt54_us.meta.llama3-3-70b-instruct-v1:0_nfp_run1 | nfp | us.meta.llama3-3-70b-instruct-v1:0 | 150 / 150 | usable | no record-level integrity findings | 3a4c776fb4c6… |
| Prompt strategy | gpt54_fewshot_gpt-5.4_fp_run1 | fp | gpt-5.4 | 585 / 585 | usable | no record-level integrity findings | ca2905c43b6c… |
| Prompt strategy | gpt54_fewshot_gpt-5.4_nfp_run1 | nfp | gpt-5.4 | 150 / 150 | usable | no record-level integrity findings | 7b90bd6fff7b… |
| Skill portability | gpt54_gpt4o_skill_gpt-5.4_fp_run1 | fp | gpt-5.4 | 585 / 585 | usable | no record-level integrity findings | 1462bfe32496… |
| Skill portability | gpt54_gpt4o_skill_gpt-5.4_nfp_run1 | nfp | gpt-5.4 | 150 / 150 | usable | no record-level integrity findings | 0063d7074ea3… |
| Prompt strategy | llm_2k_gemini-2.5-flash_fp_run1 | fp | gemini-2.5-flash | 585 / 557 | usable with conservative missing-as-failure analysis | invalid_scores=28 | 8674067d7eaa… |
| Prompt strategy | llm_2k_gemini-2.5-flash_nfp_run1 | nfp | gemini-2.5-flash | 150 / 147 | usable with conservative missing-as-failure analysis | invalid_scores=3 | 77a475dd7c1d… |
| Prompt strategy | llm_2k_gemini-3.5-flash_fp_run1 | fp | gemini-3.5-flash | 585 / 584 | usable with conservative missing-as-failure analysis | target_errors=1; judge_errors=1; blank_responses=1; invalid_scores=1 | 7eb9d8369590… |
| Prompt strategy | llm_2k_gemini-3.5-flash_nfp_run1 | nfp | gemini-3.5-flash | 150 / 150 | usable | no record-level integrity findings | 89f487bf20e6… |
| Prompt strategy | llm_2k_gpt-4o-2024-11-20_fp_run1 | fp | gpt-4o-2024-11-20 | 585 / 585 | usable | no record-level integrity findings | 6b67d1add93d… |
| Prompt strategy | llm_2k_gpt-4o-2024-11-20_nfp_run1 | nfp | gpt-4o-2024-11-20 | 150 / 150 | usable | no record-level integrity findings | 56de14fead06… |
| Prompt strategy | llm_2k_gpt-5.4_fp_run1 | fp | gpt-5.4 | 585 / 585 | usable | no record-level integrity findings | b971b840b78d… |
| Prompt strategy | llm_2k_gpt-5.4_nfp_run1 | nfp | gpt-5.4 | 150 / 150 | usable | no record-level integrity findings | 96d323111c57… |
| Prompt strategy | llm_2k_gpt-5.6-luna_fp_run1 | fp | gpt-5.6-luna | 585 / 585 | usable | no record-level integrity findings | 54f4a55d0463… |
| Prompt strategy | llm_2k_gpt-5.6-luna_nfp_run1 | nfp | gpt-5.6-luna | 150 / 150 | usable | no record-level integrity findings | e7cf2b668cab… |
| Prompt strategy | llm_2k_us.meta.llama3-1-8b-instruct-v1:0_fp_run1 | fp | us.meta.llama3-1-8b-instruct-v1:0 | 585 / 585 | usable | no record-level integrity findings | 346e3e595063… |
| Prompt strategy | llm_2k_us.meta.llama3-1-8b-instruct-v1:0_nfp_run1 | nfp | us.meta.llama3-1-8b-instruct-v1:0 | 150 / 150 | usable | no record-level integrity findings | dba810853c65… |
| Prompt strategy | llm_2k_us.meta.llama3-3-70b-instruct-v1:0_fp_run1 | fp | us.meta.llama3-3-70b-instruct-v1:0 | 585 / 585 | usable | no record-level integrity findings | da21951a68ab… |
| Prompt strategy | llm_2k_us.meta.llama3-3-70b-instruct-v1:0_nfp_run1 | nfp | us.meta.llama3-3-70b-instruct-v1:0 | 150 / 150 | usable | no record-level integrity findings | c71bc7fe2301… |
| Prompt strategy | llm_5k_gemini-2.5-flash_fp_run1 | fp | gemini-2.5-flash | 585 / 550 | usable with conservative missing-as-failure analysis | invalid_scores=35 | 4621a8e54f5d… |
| Prompt strategy | llm_5k_gemini-2.5-flash_nfp_run1 | nfp | gemini-2.5-flash | 150 / 149 | usable with conservative missing-as-failure analysis | invalid_scores=1 | c7581cd57b47… |
| Prompt strategy | llm_5k_gemini-3.5-flash_fp_run1 | fp | gemini-3.5-flash | 585 / 585 | usable | no record-level integrity findings | 21b20cfb9a4b… |
| Prompt strategy | llm_5k_gemini-3.5-flash_nfp_run1 | nfp | gemini-3.5-flash | 150 / 150 | usable | no record-level integrity findings | d2a8898ef47f… |
| Prompt strategy | llm_5k_gpt-4o-2024-11-20_fp_run1 | fp | gpt-4o-2024-11-20 | 585 / 585 | usable | no record-level integrity findings | 11f8497cbbf0… |
| Prompt strategy | llm_5k_gpt-4o-2024-11-20_nfp_run1 | nfp | gpt-4o-2024-11-20 | 150 / 150 | usable | no record-level integrity findings | 8992dd04c16e… |
| Prompt strategy | llm_5k_gpt-5.4_fp_run1 | fp | gpt-5.4 | 585 / 585 | usable | no record-level integrity findings | 0e10c1c7e89a… |
| Prompt strategy | llm_5k_gpt-5.4_nfp_run1 | nfp | gpt-5.4 | 150 / 150 | usable | no record-level integrity findings | a50b5f188ead… |
| Prompt strategy | llm_5k_gpt-5.6-luna_fp_run1 | fp | gpt-5.6-luna | 585 / 585 | usable | no record-level integrity findings | 63f3cdfdba61… |
| Prompt strategy | llm_5k_gpt-5.6-luna_nfp_run1 | nfp | gpt-5.6-luna | 150 / 150 | usable | no record-level integrity findings | 924ff1b25dd5… |
| Prompt strategy | llm_5k_us.meta.llama3-1-8b-instruct-v1:0_fp_run1 | fp | us.meta.llama3-1-8b-instruct-v1:0 | 585 / 585 | usable | no record-level integrity findings | 88bcf25bdf32… |
| Prompt strategy | llm_5k_us.meta.llama3-1-8b-instruct-v1:0_nfp_run1 | nfp | us.meta.llama3-1-8b-instruct-v1:0 | 150 / 150 | usable | no record-level integrity findings | 03cbe502a2ab… |
| Prompt strategy | llm_5k_us.meta.llama3-3-70b-instruct-v1:0_fp_run1 | fp | us.meta.llama3-3-70b-instruct-v1:0 | 585 / 585 | usable | no record-level integrity findings | 6199e221d9c7… |
| Prompt strategy | llm_5k_us.meta.llama3-3-70b-instruct-v1:0_nfp_run1 | nfp | us.meta.llama3-3-70b-instruct-v1:0 | 150 / 150 | usable | no record-level integrity findings | 7b8eaded64df… |
| Prompt strategy | llm_natural_gemini-2.5-flash_fp_run1 | fp | gemini-2.5-flash | 585 / 539 | usable with conservative missing-as-failure analysis | invalid_scores=46 | 80f11553b880… |
| Prompt strategy | llm_natural_gemini-2.5-flash_nfp_run1 | nfp | gemini-2.5-flash | 150 / 148 | usable with conservative missing-as-failure analysis | invalid_scores=2 | f5bc34131f25… |
| Prompt strategy | llm_natural_gemini-3.5-flash_fp_run1 | fp | gemini-3.5-flash | 585 / 584 | usable with conservative missing-as-failure analysis | target_errors=1; judge_errors=1; blank_responses=1; invalid_scores=1 | ed0d1e4f2325… |

| Family | Condition | Type | Model | Rows / scored | Status | Finding | SHA-256 |
|---|---|---|---|---|---|---|---|
| Prompt strategy | llm_natural_gemini-3.5-flash_nfp_run1 | nfp | gemini-3.5-flash | 150 / 150 | usable | no record-level integrity findings | 7e27259fec34… |
| Prompt strategy | llm_natural_gpt-4o-2024-11-20_fp_run1 | fp | gpt-4o-2024-11-20 | 585 / 585 | usable | no record-level integrity findings | 8e55ce64f43f… |
| Prompt strategy | llm_natural_gpt-4o-2024-11-20_nfp_run1 | nfp | gpt-4o-2024-11-20 | 150 / 150 | usable | no record-level integrity findings | 996891cc2804… |
| Prompt strategy | llm_natural_gpt-5.4_fp_run1 | fp | gpt-5.4 | 585 / 585 | usable | no record-level integrity findings | e3143cae13f6… |
| Prompt strategy | llm_natural_gpt-5.4_nfp_run1 | nfp | gpt-5.4 | 150 / 150 | usable | no record-level integrity findings | 9ef495ff7465… |
| Prompt strategy | llm_natural_gpt-5.6-luna_fp_run1 | fp | gpt-5.6-luna | 585 / 585 | usable | no record-level integrity findings | df8ddc9b3101… |
| Prompt strategy | llm_natural_gpt-5.6-luna_nfp_run1 | nfp | gpt-5.6-luna | 150 / 150 | usable | no record-level integrity findings | cd240628a0fa… |
| Prompt strategy | llm_natural_us.meta.llama3-1-8b-instruct-v1:0_fp_run1 | fp | us.meta.llama3-1-8b-instruct-v1:0 | 585 / 585 | usable | no record-level integrity findings | 55af452329b9… |
| Prompt strategy | llm_natural_us.meta.llama3-1-8b-instruct-v1:0_nfp_run1 | nfp | us.meta.llama3-1-8b-instruct-v1:0 | 150 / 150 | usable | no record-level integrity findings | 80ab5a7ec95a… |
| Prompt strategy | llm_natural_us.meta.llama3-3-70b-instruct-v1:0_fp_run1 | fp | us.meta.llama3-3-70b-instruct-v1:0 | 585 / 585 | usable | no record-level integrity findings | 14354dcc85cf… |
| Prompt strategy | llm_natural_us.meta.llama3-3-70b-instruct-v1:0_nfp_run1 | nfp | us.meta.llama3-3-70b-instruct-v1:0 | 150 / 150 | usable | no record-level integrity findings | e90683c110ba… |
| Prompt strategy | luna_fewshot_gpt-5.6-luna_fp_run1 | fp | gpt-5.6-luna | 585 / 585 | usable | no record-level integrity findings | a703467e4108… |
| Prompt strategy | luna_fewshot_gpt-5.6-luna_nfp_run1 | nfp | gpt-5.6-luna | 150 / 150 | usable | no record-level integrity findings | cf835e1241e6… |
| Skill portability | luna_gpt4o_skill_gpt-5.6-luna_fp_run1 | fp | gpt-5.6-luna | 585 / 585 | usable | no record-level integrity findings | 70dfe4854f85… |
| Skill portability | luna_gpt4o_skill_gpt-5.6-luna_nfp_run1 | nfp | gpt-5.6-luna | 150 / 150 | usable | no record-level integrity findings | 3a2021ba038a… |
| Skill portability | luna_opt_skill_gpt-4o-2024-11-20_nfp_run1 | nfp | gpt-4o-2024-11-20 | 150 / 150 | usable | no record-level integrity findings | 5ff4c4ead916… |
| Skill portability | port_gemini_skill_gpt-4o-2024-11-20_fp_run1 | fp | gpt-4o-2024-11-20 | 585 / 585 | usable | no record-level integrity findings | f755d2daea5c… |
| Skill portability | port_gemini_skill_gpt-4o-2024-11-20_nfp_run1 | nfp | gpt-4o-2024-11-20 | 150 / 150 | usable | no record-level integrity findings | 9bf1556b5a5b… |
| Skill portability | port_gpt4o_skill_gemini-2.5-flash_fp_run1 | fp | gemini-2.5-flash | 585 / 552 | usable with conservative missing-as-failure analysis | invalid_scores=33 | 00c2f9e32413… |
| Skill portability | port_gpt4o_skill_gemini-2.5-flash_nfp_run1 | nfp | gemini-2.5-flash | 150 / 148 | usable with conservative missing-as-failure analysis | invalid_scores=2 | c4a94c6c4668… |
| Skill portability | port_gpt4o_skill_gemini-3.5-flash_fp_run1 | fp | gemini-3.5-flash | 585 / 585 | usable | no record-level integrity findings | abdb11c1e3db… |
| Skill portability | port_gpt4o_skill_gemini-3.5-flash_nfp_run1 | nfp | gemini-3.5-flash | 150 / 150 | usable | no record-level integrity findings | 4779ae53c6b0… |
| Skill portability | port_gpt4o_skill_us.meta.llama3-1-8b-instruct-v1:0_fp_run1 | fp | us.meta.llama3-1-8b-instruct-v1:0 | 585 / 585 | usable | no record-level integrity findings | 633c177fe2c7… |
| Skill portability | port_gpt4o_skill_us.meta.llama3-1-8b-instruct-v1:0_nfp_run1 | nfp | us.meta.llama3-1-8b-instruct-v1:0 | 150 / 150 | usable | no record-level integrity findings | b591a138801e… |
| Skill portability | port_gpt4o_skill_us.meta.llama3-3-70b-instruct-v1:0_fp_run1 | fp | us.meta.llama3-3-70b-instruct-v1:0 | 585 / 585 | usable | no record-level integrity findings | f66cf14fe0ee… |
| Skill portability | port_gpt4o_skill_us.meta.llama3-3-70b-instruct-v1:0_nfp_run1 | nfp | us.meta.llama3-3-70b-instruct-v1:0 | 150 / 150 | usable | no record-level integrity findings | 801c3e392462… |
| Skill portability | port_gpt54_skill_gemini-2.5-flash_fp_run1 | fp | gemini-2.5-flash | 585 / 551 | usable with conservative missing-as-failure analysis | invalid_scores=34 | a7615ae66c76… |
| Skill portability | port_gpt54_skill_gemini-2.5-flash_nfp_run1 | nfp | gemini-2.5-flash | 150 / 150 | usable | no record-level integrity findings | f97c1c8d182e… |
| Skill portability | port_gpt54_skill_gpt-4o-2024-11-20_fp_run1 | fp | gpt-4o-2024-11-20 | 585 / 585 | usable | no record-level integrity findings | 8afceb3cb44e… |
| Skill portability | port_gpt54_skill_gpt-4o-2024-11-20_nfp_run1 | nfp | gpt-4o-2024-11-20 | 150 / 150 | usable | no record-level integrity findings | 4caf265a825b… |
| Skill portability | port_rep1_skill_gemini-3.5-flash_fp_run1 | fp | gemini-3.5-flash | 585 / 585 | usable | no record-level integrity findings | 41441569f8f5… |
| Skill portability | port_rep1_skill_gpt-5.4_fp_run1 | fp | gpt-5.4 | 585 / 584 | usable with conservative missing-as-failure analysis | target_errors=1; judge_errors=1; blank_responses=1; invalid_scores=1 | fa25140e6b92… |
| Skill portability | port_rep1_skill_us.meta.llama3-3-70b-instruct-v1:0_fp_run1 | fp | us.meta.llama3-3-70b-instruct-v1:0 | 585 / 585 | usable | no record-level integrity findings | 595a238dd739… |
| Skill portability | port_rep2_skill_gemini-3.5-flash_fp_run1 | fp | gemini-3.5-flash | 585 / 585 | usable | no record-level integrity findings | c6305b51afa7… |
| Skill portability | port_rep2_skill_gpt-5.4_fp_run1 | fp | gpt-5.4 | 585 / 584 | usable with conservative missing-as-failure analysis | target_errors=1; judge_errors=1; blank_responses=1; invalid_scores=1 | 55537ab946c4… |
| Skill portability | port_rep2_skill_us.meta.llama3-3-70b-instruct-v1:0_fp_run1 | fp | us.meta.llama3-3-70b-instruct-v1:0 | 585 / 585 | usable | no record-level integrity findings | 752a4a546562… |
| Legacy optimisation | skillopt_v0001_gpt-4o-2024-11-20_fp_run1 | fp | gpt-4o-2024-11-20 | 585 / 585 | usable | no record-level integrity findings | d50236cd0021… |
| Legacy optimisation | skillopt_v0001_gpt-4o-2024-11-20_nfp_run1 | nfp | gpt-4o-2024-11-20 | 150 / 150 | usable | no record-level integrity findings | cc8de53d6005… |
| Skill evaluation | skillopt_v2_best_llama-3.1-8b_nfp_run1 | nfp | llama-3.1-8b | 150 / 150 | usable | no record-level integrity findings | 08311f711919… |

| Family | Condition | Type | Model | Rows / scored | Status | Finding | SHA-256 |
|---|---|---|---|---|---|---|---|
| Skill evaluation | skillopt_v2_gemini_best_gemini-2.5-flash_nfp_run1 | nfp | gemini-2.5-flash | 150 / 150 | usable | no record-level integrity findings | 0edb1ec3ba1f… |
| Skill evaluation | skillopt_v2_gpt4o_best_gpt-4o-2024-11-20_fp_run1 | fp | gpt-4o-2024-11-20 | 585 / 585 | usable | no record-level integrity findings | 6bc57dd2595e… |
| Skill evaluation | skillopt_v2_gpt4o_best_gpt-4o-2024-11-20_nfp_run1 | nfp | gpt-4o-2024-11-20 | 150 / 150 | usable | no record-level integrity findings | a5746f04348a… |
| Skill evaluation | skillopt_v2_gpt54_best_gpt-5.4_nfp_run1 | nfp | gpt-5.4 | 150 / 150 | usable | no record-level integrity findings | a9a304b9f14e… |

#### S2. Dataset mapping and analysis populations

The original Cancer-Myth source uses sparse QIDs. The external evaluation harness uses sequential row numbers. We verified that the 585 records in the source and flattened corpus matched exactly by QID and question text, then mapped the 410 saved test QIDs to their sequential external IDs.

The NFP dataset contained 150 records. The 30 NFP questions used for validation were reconstructed from the recorded loader seed; the remaining 120 formed the NFP evaluation set. All cross-model FP and NFP analyses used these common 410 and 120-question populations.

##### Table S2. Primary source registry and hashes

| Run | Condition | Rows | Result SHA-256 | Summary SHA-256 |
|---|---|---|---|---|
| skillopt_v2_gpt4o | Baseline | 410 | b2dc0f6115e6… | 5324f033aea6… |
| skillopt_v2_gpt4o | Post-optimisation | 410 | 361e11fb6c0f… | 3667965963ad… |
| skillopt_v2_gpt4o_replicate1 | Baseline | 410 | 563d62889e3b… | 0ff936d98255… |
| skillopt_v2_gpt4o_replicate1 | Post-optimisation | 410 | a3ec812eed98… | 0cf351c08de4… |
| skillopt_v2_gpt4o_replicate2 | Baseline | 410 | 3c84b29c536f… | e9901be0bec5… |
| skillopt_v2_gpt4o_replicate2 | Post-optimisation | 410 | ccbea2e4e019… | df27d59e037b… |

#### S3. SkillOpt configuration

All three primary executions used GPT-4o-2024-11-20 as target and GPT-5.4-2026-03-05 as optimiser. The data split and recorded seed were 42. Each execution began with the same 128-character instruction and had 12 planned steps, with batches of 40 FP questions. In rewrite_from_suggestions mode, scored outputs from successes and failures were aggregated and supplied to the rewrite model. Full-skill rewrites allowed 2,000 completion tokens, with an initial edit budget of four and minimum of two. Candidate skills were selected using one combined hard-accuracy score across 58 FP and 30 NFP validation questions; no separate threshold was imposed for either outcome.

The compact run used the same process with a 1,000-token rewrite limit. Its 1,969-character step-11 checkpoint was used in the cross-model compact comparison; a distinct 2,874-character skill accepted at step 14 was used for the final compact-run evaluation. Execution 1 was continued from 12 to 24 steps as an exploratory convergence check; its retained step-3 skill did not change. Executions 2 and 3 ended after the planned 12 steps.

##### Table S3. SkillOpt training runs, settings and completion status

| Experiment | Target | Optimiser | Steps | Accepted | Baseline | Final | Change | Skill chars | Status |
|---|---|---|---|---|---|---|---|---|---|
| GPT-4o / GPT-5.4 optimiser (execution 1) | gpt-4o-2024-11-20 | gpt-5.4 | 24/24 | 2 | 16.3 | 82.7 | 66.3 | 4933 | complete paired evaluation |
| GPT-4o / GPT-5.4 optimiser (execution 2) | gpt-4o-2024-11-20 | gpt-5.4 | 12/12 | 3 | 17.6 | 90.5 | 72.9 | 4414 | complete paired evaluation |
| GPT-4o / GPT-5.4 optimiser (execution 3) | gpt-4o-2024-11-20 | gpt-5.4 | 12/12 | 2 | 15.4 | 82.2 | 66.8 | 5416 | complete paired evaluation |
| GPT-4o / GPT-5.4 optimiser (compact) | gpt-4o-2024-11-20 | gpt-5.4 | 24/24 | 2 | 15.1 | 61.2 | 46.1 | 2874 | complete paired evaluation |
| GPT-4o self-optimisation | gpt-4o-2024-11-20 | gpt-4o-2024-11-20 | 12/12 | 0 | 16.8 | – | – | 128 | complete optimisation; no learned rewrite |
| GPT-4o / GPT-5.6 Luna optimiser | gpt-4o-2024-11-20 | gpt-5.6-luna | 12/12 | 3 | 17.6 | 58.8 | 41.2 | 7051 | complete paired evaluation |

| Experiment | Target | Optimiser | Steps | Accepted | Baseline | Final | Change | Skill chars | Status |
|---|---|---|---|---|---|---|---|---|---|
| GPT-5.4 self-optimisation | gpt-5.4-2026-03-05 | gpt-5.4 | 12/12 | 2 | 59.5 | 89.8 | 30.2 | 5352 | complete paired evaluation |
| Gemini 2.5 Flash / GPT-5.4 optimiser | gemini-2.5-flash | gpt-5.4-2026-03-05 | 12/12 | 3 | 2.0 | 20.5 | 18.5 | 4806 | complete paired evaluation |
| Gemini 3.5 Flash / GPT-5.4 optimiser | gemini-3.5-flash | gpt-5.4 | 0/12 | 0 | – | – | – | – | not started |
| Llama 3.1 8B / GPT-5.4 optimiser | us.meta.llama3-1-8b-instruct-v1:0 | gpt-5.4 | 3/12 | 0 | – | – | – | – | incomplete optimisation |
| Llama 3.3 70B / GPT-5.4 optimiser | us.meta.llama3-3-70b-instruct-v1:0 | gpt-5.4 | 4/12 | 0 | – | – | – | – | incomplete optimisation |
| GPT-5.6 Luna / GPT-5.4 optimiser | gpt-5.6-luna | gpt-5.4 | 0/12 | 0 | – | – | – | – | not started |

### S4. Primary GPT-4o analysis

FP correction was the proportion of responses assigned +1 by the Cancer-Myth judge. Each comparison used the same 410 questions. Wilson intervals were calculated for condition-specific proportions. Absolute differences and their 95% confidence intervals used 10,000 paired bootstrap samples. Exact two-sided McNemar tests compared discordant pairs and were adjusted across the three runs using Holm's method.

#### Table S4. Primary condition-specific estimates

| Run | Condition | Success / N | % (95% CI) |
|---|---|---|---|
| skillopt_v2_gpt4o | Baseline | 67 / 410 | 16.3 (13.1–20.2) |
| skillopt_v2_gpt4o | Post-optimisation | 339 / 410 | 82.7 (78.7–86.0) |
| skillopt_v2_gpt4o_replicate1 | Baseline | 72 / 410 | 17.6 (14.2–21.5) |
| skillopt_v2_gpt4o_replicate1 | Post-optimisation | 371 / 410 | 90.5 (87.3–93.0) |
| skillopt_v2_gpt4o_replicate2 | Baseline | 63 / 410 | 15.4 (12.2–19.2) |
| skillopt_v2_gpt4o_replicate2 | Post-optimisation | 337 / 410 | 82.2 (78.2–85.6) |

#### Table S5. Paired primary comparisons

| Run | Baseline, % | SkillOpt, % | Difference, pp (95% CI) | Improved / worsened | Holm p |
|---|---|---|---|---|---|
| 1 | 16.3 | 82.7 | 66.3 (61.7 to 71.0) | 273 / 1 | 1.8e-80 |
| 2 | 17.6 | 90.5 | 72.9 (68.5 to 77.3) | 299 / 0 | 5.9e-90 |
| 3 | 15.4 | 82.2 | 66.8 (62.2 to 71.2) | 274 / 0 | 1.3e-82 |

#### Table S6. Category-level correction estimates

| Category | Run | N | Baseline % | SkillOpt % | Difference, pp (95% CI) |
|---|---|---|---|---|---|
| causal misattribution | skillopt_v2_gpt4o | 49 | 28.6 | 89.8 | 61.2 (46.9 to 75.5) |
| causal misattribution | skillopt_v2_gpt4o_replicate1 | 49 | 34.7 | 98.0 | 63.3 (49.0 to 77.6) |
| causal misattribution | skillopt_v2_gpt4o_replicate2 | 49 | 20.4 | 91.8 | 71.4 (59.2 to 83.7) |
| inevitable side effect | skillopt_v2_gpt4o | 78 | 7.7 | 85.9 | 78.2 (69.2 to 87.2) |
| inevitable side effect | skillopt_v2_gpt4o_replicate1 | 78 | 10.3 | 93.6 | 83.3 (74.4 to 91.0) |
| inevitable side effect | skillopt_v2_gpt4o_replicate2 | 78 | 11.5 | 87.2 | 75.6 (65.4 to 84.6) |
| no symptoms means no disease | skillopt_v2_gpt4o | 33 | 24.2 | 87.9 | 63.6 (45.5 to 78.8) |
| no symptoms means no disease | skillopt_v2_gpt4o_replicate1 | 33 | 15.2 | 84.8 | 69.7 (54.5 to 84.8) |
| no symptoms means no disease | skillopt_v2_gpt4o_replicate2 | 33 | 30.3 | 81.8 | 51.5 (33.3 to 69.7) |
| no treatment | skillopt_v2_gpt4o | 89 | 14.6 | 85.4 | 70.8 (60.7 to 79.8) |
| no treatment | skillopt_v2_gpt4o_replicate1 | 89 | 12.4 | 94.4 | 82.0 (74.2 to 89.9) |
| no treatment | skillopt_v2_gpt4o_replicate2 | 89 | 7.9 | 87.6 | 79.8 (70.8 to 87.6) |
| only/standard treatment | skillopt_v2_gpt4o | 90 | 13.3 | 77.8 | 64.4 (54.4 to 74.4) |
| only/standard treatment | skillopt_v2_gpt4o_replicate1 | 90 | 16.7 | 86.7 | 70.0 (60.0 to 78.9) |
| only/standard treatment | skillopt_v2_gpt4o_replicate2 | 90 | 13.3 | 77.8 | 64.4 (54.4 to 74.4) |
| other | skillopt_v2_gpt4o | 26 | 26.9 | 76.9 | 50.0 (30.8 to 69.2) |
| other | skillopt_v2_gpt4o_replicate1 | 26 | 26.9 | 84.6 | 57.7 (38.5 to 76.9) |

| Category | Run | N | Baseline % | SkillOpt % | Difference, pp (95% CI) |
|---|---|---|---|---|---|
| other | skillopt_v2_gpt4o_replicate2 | 26 | 19.2 | 65.4 | 46.2 (26.9 to 65.4) |
| underestimate risk | skillopt_v2_gpt4o | 45 | 15.6 | 73.3 | 57.8 (44.4 to 71.1) |
| underestimate risk | skillopt_v2_gpt4o_replicate1 | 45 | 20.0 | 84.4 | 64.4 (51.1 to 77.8) |
| underestimate risk | skillopt_v2_gpt4o_replicate2 | 45 | 22.2 | 71.1 | 48.9 (33.3 to 64.4) |

##### Table S7. Cross-run question consistency

| Successful baseline runs | Successful SkillOpt runs | Questions |
|---|---|---|
| 0 | 0 | 26 |
| 0 | 1 | 28 |
| 0 | 2 | 48 |
| 0 | 3 | 211 |
| 1 | 0 | 0 |
| 1 | 1 | 0 |
| 1 | 2 | 1 |
| 1 | 3 | 31 |
| 2 | 0 | 0 |
| 2 | 1 | 0 |
| 2 | 2 | 0 |
| 2 | 3 | 25 |
| 3 | 0 | 0 |
| 3 | 1 | 0 |
| 3 | 2 | 0 |
| 3 | 3 | 40 |

#### S5. Prompt and skill provenance

The deliberately strong few-shot comparator was generated once by GPT-5.4-2026-03-05 at temperature 0 and was blinded to SkillOpt outputs. GPT-5.4 selected five FP examples from 30 candidates in the training split and two NFP examples from 20 candidate controls, then wrote exemplar responses. The resulting prompt contained seven worked examples and measured 5,927 characters. The instruction-only comparators were also generated once by GPT-5.4 at temperature 0 using 30 FP training examples as task context. The same template generated an unconstrained prompt and prompts targeting approximately 5,000 and 2,000 characters. Final lengths were 14,533, 5,774 and 2,814 characters; none contained worked examples. Bespoke human-written prompts were excluded because author wording would introduce an uncontrolled comparator that could not be standardised or reproduced.

The external harness passed each complete prompt to the target model but stored only its first 200 characters in the response ledger. Prompt identity was reconstructed from condition labels, matching prefixes, surviving full artifacts and experiment chronology. All manuscript prompt groups resolved to a surviving full artifact. The external compact comparison and the final compact-run evaluation used the distinct skill versions described above and are not treated as the same intervention.

##### Table S8. Prompt and skill artifact registry

| Artifact | Path | Characters | Words | SHA-256 | Provenance |
|---|---|---|---|---|---|
| Natural rule prompt | skills/llm_instructions_natural.md | 14533 | 2152 | 26ddcf569c21… | one-shot GPT-5.4 instruction generation |
| Medium rule prompt | skills/llm_instructions_5k.md | 5774 | 846 | 56e349b7c6d6… | one-shot GPT-5.4 instruction generation |
| Short rule prompt | skills/llm_instructions_2k.md | 2814 | 410 | b45fae07045c… | one-shot GPT-5.4 instruction generation |
| Few-shot prompt | skills/fewshot_gpt54.md | 5927 | 929 | 88448e444457… | one-shot GPT-5.4 prompt with worked examples |
| Primary learned skill | results/skillopt_v2_gpt4o/best_skill.md | 4933 | 703 | 7b2d733d2736… | SkillOpt execution 1 |

| Artifact | Path | Characters | Words | SHA-256 | Provenance |
|---|---|---|---|---|---|
| Learned skill replicate 1 | results/skillopt_v2_gpt4o_replicate1/best_skill.md | 4414 | 626 | 98cc6816a960… | SkillOpt execution 2 |
| Learned skill replicate 2 | results/skillopt_v2_gpt4o_replicate2/best_skill.md | 5416 | 779 | 83d8d5f5366b… | SkillOpt execution 3 |
| Compact checkpoint | results/skillopt_v2_gpt4o_compact/skills/skill_v0011.md | 1969 | 274 | 4fda388ff50b… | SkillOpt compact step 11 used in external grid |
| Compact final skill | results/skillopt_v2_gpt4o_compact/best_skill.md | 2874 | 423 | 34dcd71d7a9c… | SkillOpt compact step 14 native best |

### Table S9. Prompt-lineage registry

| Condition | Model | Type | Full artifact | Chars | Lineage | Ledger SHA-256 |
|---|---|---|---|---|---|---|
| comp_gemini_gemini-2.5-flash_fp_run1 | gemini-2.5-flash | fp | skills/composite_gpt4o.md or skills/composite_gemini_flash.md | – | resolved by composite condition label; exact component file is model-specific | 9647edb05fe3… |
| comp_gemini_gemini-2.5-flash_nfp_run1 | gemini-2.5-flash | nfp | skills/composite_gpt4o.md or skills/composite_gemini_flash.md | – | resolved by composite condition label; exact component file is model-specific | 61272d3e77e0… |
| comp_gpt4o_gpt-4o-2024-11-20_fp_run1 | gpt-4o-2024-11-20 | fp | skills/composite_gpt4o.md or skills/composite_gemini_flash.md | – | resolved by composite condition label; exact component file is model-specific | 9f30cadb064e… |
| comp_gpt4o_gpt-4o-2024-11-20_nfp_run1 | gpt-4o-2024-11-20 | nfp | skills/composite_gpt4o.md or skills/composite_gemini_flash.md | – | resolved by composite condition label; exact component file is model-specific | df4cb448acbe… |
| compact_skill_gemini-2.5-flash_fp_run1 | gemini-2.5-flash | fp | results/skillopt_v2_gpt4o_compact/skills/skill_v0011.md | 1969 | resolved | 43c94262632c… |
| compact_skill_gemini-2.5-flash_nfp_run1 | gemini-2.5-flash | nfp | results/skillopt_v2_gpt4o_compact/skills/skill_v0011.md | 1969 | resolved | cfc8815a2e75… |
| compact_skill_gemini-3.5-flash_fp_run1 | gemini-3.5-flash | fp | results/skillopt_v2_gpt4o_compact/skills/skill_v0011.md | 1969 | resolved | f7718ee1e19d… |
| compact_skill_gemini-3.5-flash_nfp_run1 | gemini-3.5-flash | nfp | results/skillopt_v2_gpt4o_compact/skills/skill_v0011.md | 1969 | resolved | 9374ef5d8111… |
| compact_skill_gpt-4o-2024-11-20_fp_run1 | gpt-4o-2024-11-20 | fp | results/skillopt_v2_gpt4o_compact/skills/skill_v0011.md | 1969 | resolved | caabed18f0ef… |
| compact_skill_gpt-4o-2024-11-20_nfp_run1 | gpt-4o-2024-11-20 | nfp | results/skillopt_v2_gpt4o_compact/skills/skill_v0011.md | 1969 | resolved | d5f0eb03536e… |
| compact_skill_gpt-5.4_fp_run1 | gpt-5.4 | fp | results/skillopt_v2_gpt4o_compact/skills/skill_v0011.md | 1969 | resolved | c42482443049… |
| compact_skill_gpt-5.4_nfp_run1 | gpt-5.4 | nfp | results/skillopt_v2_gpt4o_compact/skills/skill_v0011.md | 1969 | resolved | 6d30787b4251… |
| compact_skill_gpt-5.6-luna_fp_run1 | gpt-5.6-luna | fp | results/skillopt_v2_gpt4o_compact/skills/skill_v0011.md | 1969 | resolved | 8b7d8407c860… |
| compact_skill_gpt-5.6-luna_nfp_run1 | gpt-5.6-luna | nfp | results/skillopt_v2_gpt4o_compact/skills/skill_v0011.md | 1969 | resolved | 120cde872f6c… |
| compact_skill_us.meta.llama3-1-8b-instruct-v1:0_fp_run1 | us.meta.llama3-1-8b-instruct-v1:0 | fp | results/skillopt_v2_gpt4o_compact/skills/skill_v0011.md | 1969 | resolved | 25f32442f893… |
| compact_skill_us.meta.llama3-1-8b-instruct-v1:0_nfp_run1 | us.meta.llama3-1-8b-instruct-v1:0 | nfp | results/skillopt_v2_gpt4o_compact/skills/skill_v0011.md | 1969 | resolved | c6e1009ff4cf… |
| compact_skill_us.meta.llama3-3-70b-instruct-v1:0_fp_run1 | us.meta.llama3-3-70b-instruct-v1:0 | fp | results/skillopt_v2_gpt4o_compact/skills/skill_v0011.md | 1969 | resolved | b662b5080202… |
| compact_skill_us.meta.llama3-3-70b-instruct-v1:0_nfp_run1 | us.meta.llama3-3-70b-instruct-v1:0 | nfp | results/skillopt_v2_gpt4o_compact/skills/skill_v0011.md | 1969 | resolved | f80225b9183a… |
| exp_a_baseline_gpt-4o-2024-11-20_fp_run1 | gpt-4o-2024-11-20 | fp | see matching baseline/few-shot/skill condition definition | – | resolved by condition label and shared intervention files | f3e23dc2dbb8… |
| exp_a_baseline_gpt-5.4_fp_run1 | gpt-5.4 | fp | see matching baseline/few-shot/skill condition definition | – | resolved by condition label and shared intervention files | 1f2f70707a99… |
| exp_a_baseline_us.meta.llama3-3-70b-instruct-v1:0_fp_run1 | us.meta.llama3-3-70b-instruct-v1:0 | fp | see matching baseline/few-shot/skill condition definition | – | resolved by condition label and shared intervention files | bbbd766aeb3b… |
| exp_a_fewshot_gpt-4o-2024-11-20_fp_run1 | gpt-4o-2024-11-20 | fp | see matching baseline/few-shot/skill condition definition | – | resolved by condition label and shared intervention files | 79a79de97f78… |
| exp_a_fewshot_gpt-5.4_fp_run1 | gpt-5.4 | fp | see matching baseline/few-shot/skill condition definition | – | resolved by condition label and shared intervention files | fa5717ce2de4… |
| exp_a_fewshot_us.meta.llama3-3-70b-instruct-v1:0_fp_run1 | us.meta.llama3-3-70b-instruct-v1:0 | fp | see matching baseline/few-shot/skill condition definition | – | resolved by condition label and shared intervention files | 8453198f2c8a… |
| exp_a_skillopt_gpt-4o-2024-11-20_fp_run1 | gpt-4o-2024-11-20 | fp | see matching baseline/few-shot/skill condition definition | – | resolved by condition label and shared intervention files | d1dc8948d40a… |
| exp_a_skillopt_gpt-5.4_fp_run1 | gpt-5.4 | fp | see matching baseline/few-shot/skill condition definition | – | resolved by condition label and shared intervention files | c2213048c1b4… |
| exp_a_skillopt_us.meta.llama3-3-70b-instruct-v1:0_fp_run1 | us.meta.llama3-3-70b-instruct-v1:0 | fp | see matching baseline/few-shot/skill condition definition | – | resolved by condition label and shared intervention files | 65b3e40232ff… |
| exp_b2_baseline_gpt-4o-2024-11-20_nfp_run1 | gpt-4o-2024-11-20 | nfp | see matching baseline/few-shot/skill condition definition | – | resolved by condition label and shared intervention files | 8d0c5ba0b730… |
| exp_b2_baseline_gpt-5.4_nfp_run1 | gpt-5.4 | nfp | see matching baseline/few-shot/skill condition definition | – | resolved by condition label and shared intervention files | a412d6a6859d… |
| exp_b2_baseline_us.meta.llama3-3-70b-instruct-v1:0_nfp_run1 | us.meta.llama3-3-70b-instruct-v1:0 | nfp | see matching baseline/few-shot/skill condition definition | – | resolved by condition label and shared intervention files | 8e56cc03e3dd… |
| exp_b2_fewshot_gpt-4o-2024-11-20_nfp_run1 | gpt-4o-2024-11-20 | nfp | see matching baseline/few-shot/skill condition definition | – | resolved by condition label and shared intervention files | cb9c7bf78662… |
| exp_b2_fewshot_gpt-5.4_nfp_run1 | gpt-5.4 | nfp | see matching baseline/few-shot/skill condition definition | – | resolved by condition label and shared intervention files | 3d7b6458e680… |
| exp_b2_fewshot_us.meta.llama3-3-70b-instruct-v1:0_nfp_run1 | us.meta.llama3-3-70b-instruct-v1:0 | nfp | see matching baseline/few-shot/skill condition definition | – | resolved by condition label and shared intervention files | d4e1a5dd67ac… |
| exp_b2_skillopt_gpt-4o-2024-11-20_nfp_run1 | gpt-4o-2024-11-20 | nfp | see matching baseline/few-shot/skill condition definition | – | resolved by condition label and shared intervention files | 9dff6c140d36… |
| exp_b2_skillopt_gpt-5.4_nfp_run1 | gpt-5.4 | nfp | see matching baseline/few-shot/skill condition definition | – | resolved by condition label and shared intervention files | 33b135eeb020… |

| Condition | Model | Type | Full artifact | Chars | Lineage | Ledger SHA-256 |
|---|---|---|---|---|---|---|
| exp_b2_skillopt_us.meta.llama3-3-70b-instruct-v1:0_nfp_run1 | us.meta.llama3-3-70b-instruct-v1:0 | nfp | see matching baseline/few-shot/skill condition definition | – | resolved by condition label and shared intervention files | 33c6893e1bc2… |
| exp_b_baseline_gpt-4o-2024-11-20_fp_run1 | gpt-4o-2024-11-20 | fp | see matching baseline/few-shot/skill condition definition | – | resolved by condition label and shared intervention files | 53bebad64d02… |
| exp_b_baseline_gpt-5.4_fp_run1 | gpt-5.4 | fp | see matching baseline/few-shot/skill condition definition | – | resolved by condition label and shared intervention files | 4b69a6b7d126… |
| exp_b_baseline_us.meta.llama3-3-70b-instruct-v1:0_fp_run1 | us.meta.llama3-3-70b-instruct-v1:0 | fp | see matching baseline/few-shot/skill condition definition | – | resolved by condition label and shared intervention files | 6afe36f083e0… |
| exp_b_fewshot_gpt-4o-2024-11-20_fp_run1 | gpt-4o-2024-11-20 | fp | see matching baseline/few-shot/skill condition definition | – | resolved by condition label and shared intervention files | 99c4a0664025… |
| exp_b_fewshot_gpt-5.4_fp_run1 | gpt-5.4 | fp | see matching baseline/few-shot/skill condition definition | – | resolved by condition label and shared intervention files | 372d09db9a00… |
| exp_b_fewshot_us.meta.llama3-3-70b-instruct-v1:0_fp_run1 | us.meta.llama3-3-70b-instruct-v1:0 | fp | see matching baseline/few-shot/skill condition definition | – | resolved by condition label and shared intervention files | 231e243cdc39… |
| exp_b_skillopt_gpt-4o-2024-11-20_fp_run1 | gpt-4o-2024-11-20 | fp | see matching baseline/few-shot/skill condition definition | – | resolved by condition label and shared intervention files | e197800dcd92… |
| exp_b_skillopt_gpt-5.4_fp_run1 | gpt-5.4 | fp | see matching baseline/few-shot/skill condition definition | – | resolved by condition label and shared intervention files | 6f148af91909… |
| exp_b_skillopt_us.meta.llama3-3-70b-instruct-v1:0_fp_run1 | us.meta.llama3-3-70b-instruct-v1:0 | fp | see matching baseline/few-shot/skill condition definition | – | resolved by condition label and shared intervention files | b1a45fb71f16… |
| fewshot_gpt54_gemini-2.5-flash_fp_run1 | gemini-2.5-flash | fp | skills/fewshot_gpt54.md | 5927 | resolved | 1c59172f5c30… |
| fewshot_gpt54_gemini-2.5-flash_nfp_run1 | gemini-2.5-flash | nfp | skills/fewshot_gpt54.md | 5927 | resolved | 937fa227254c… |
| fewshot_gpt54_gemini-3.5-flash_fp_run1 | gemini-3.5-flash | fp | skills/fewshot_gpt54.md | 5927 | resolved | 93a878ce53a8… |
| fewshot_gpt54_gemini-3.5-flash_nfp_run1 | gemini-3.5-flash | nfp | skills/fewshot_gpt54.md | 5927 | resolved | 03613e4c53fe… |
| fewshot_gpt54_gpt-4o-2024-11-20_fp_run1 | gpt-4o-2024-11-20 | fp | skills/fewshot_gpt54.md | 5927 | resolved | ea9fc5a5e8d7… |
| fewshot_gpt54_gpt-4o-2024-11-20_nfp_run1 | gpt-4o-2024-11-20 | nfp | skills/fewshot_gpt54.md | 5927 | resolved | 5d921f4e9498… |
| fewshot_gpt54_llama-3.1-8b_fp_run1 | llama-3.1-8b | fp | skills/fewshot_gpt54.md | 5927 | resolved | 2038292e2c69… |
| fewshot_gpt54_llama-3.1-8b_nfp_run1 | llama-3.1-8b | nfp | skills/fewshot_gpt54.md | 5927 | resolved | c70fcbdef35f… |
| fewshot_gpt54_us.meta.llama3-1-8b-instruct-v1:0_fp_run1 | us.meta.llama3-1-8b-instruct-v1:0 | fp | skills/fewshot_gpt54.md | 5927 | resolved | 3d3cb44af552… |
| fewshot_gpt54_us.meta.llama3-1-8b-instruct-v1:0_nfp_run1 | us.meta.llama3-1-8b-instruct-v1:0 | nfp | skills/fewshot_gpt54.md | 5927 | resolved | 9e083dde0987… |
| fewshot_gpt54_us.meta.llama3-3-70b-instruct-v1:0_fp_run1 | us.meta.llama3-3-70b-instruct-v1:0 | fp | skills/fewshot_gpt54.md | 5927 | resolved | 4e0f52ca326c… |
| fewshot_gpt54_us.meta.llama3-3-70b-instruct-v1:0_nfp_run1 | us.meta.llama3-3-70b-instruct-v1:0 | nfp | skills/fewshot_gpt54.md | 5927 | resolved | 3a4c776fb4c6… |
| gpt54_fewshot_gpt-5.4_fp_run1 | gpt-5.4 | fp | skills/fewshot_gpt54.md | 5927 | resolved | ca2905c43b6c… |
| gpt54_fewshot_gpt-5.4_nfp_run1 | gpt-5.4 | nfp | skills/fewshot_gpt54.md | 5927 | resolved | 7b90bd6fff7b… |
| gpt54_gpt4o_skill_gpt-5.4_fp_run1 | gpt-5.4 | fp | results/skillopt_v2_gpt4o/best_skill.md | 4933 | resolved | 1462bfe32496… |
| gpt54_gpt4o_skill_gpt-5.4_nfp_run1 | gpt-5.4 | nfp | results/skillopt_v2_gpt4o/best_skill.md | 4933 | resolved | 0063d7074ea3… |
| llm_2k_gemini-2.5-flash_fp_run1 | gemini-2.5-flash | fp | skills/llm_instructions_2k.md | 2814 | resolved | 8674067d7eaa… |
| llm_2k_gemini-2.5-flash_nfp_run1 | gemini-2.5-flash | nfp | skills/llm_instructions_2k.md | 2814 | resolved | 77a475dd7c1d… |
| llm_2k_gemini-3.5-flash_fp_run1 | gemini-3.5-flash | fp | skills/llm_instructions_2k.md | 2814 | resolved | 7eb9d8369590… |
| llm_2k_gemini-3.5-flash_nfp_run1 | gemini-3.5-flash | nfp | skills/llm_instructions_2k.md | 2814 | resolved | 89f487bf20e6… |
| llm_2k_gpt-4o-2024-11-20_fp_run1 | gpt-4o-2024-11-20 | fp | skills/llm_instructions_2k.md | 2814 | resolved | 6b67d1add93d… |
| llm_2k_gpt-4o-2024-11-20_nfp_run1 | gpt-4o-2024-11-20 | nfp | skills/llm_instructions_2k.md | 2814 | resolved | 56de14fead06… |
| llm_2k_gpt-5.4_fp_run1 | gpt-5.4 | fp | skills/llm_instructions_2k.md | 2814 | resolved | b971b840b78d… |
| llm_2k_gpt-5.4_nfp_run1 | gpt-5.4 | nfp | skills/llm_instructions_2k.md | 2814 | resolved | 96d323111c57… |
| llm_2k_gpt-5.6-luna_fp_run1 | gpt-5.6-luna | fp | skills/llm_instructions_2k.md | 2814 | resolved | 54f4a55d0463… |
| llm_2k_gpt-5.6-luna_nfp_run1 | gpt-5.6-luna | nfp | skills/llm_instructions_2k.md | 2814 | resolved | e7cf2b668cab… |
| llm_2k_us.meta.llama3-1-8b-instruct-v1:0_fp_run1 | us.meta.llama3-1-8b-instruct-v1:0 | fp | skills/llm_instructions_2k.md | 2814 | resolved | 346e3e595063… |
| llm_2k_us.meta.llama3-1-8b-instruct-v1:0_nfp_run1 | us.meta.llama3-1-8b-instruct-v1:0 | nfp | skills/llm_instructions_2k.md | 2814 | resolved | dba810853c65… |
| llm_2k_us.meta.llama3-3-70b-instruct-v1:0_fp_run1 | us.meta.llama3-3-70b-instruct-v1:0 | fp | skills/llm_instructions_2k.md | 2814 | resolved | da21951a68ab… |
| llm_2k_us.meta.llama3-3-70b-instruct-v1:0_nfp_run1 | us.meta.llama3-3-70b-instruct-v1:0 | nfp | skills/llm_instructions_2k.md | 2814 | resolved | c71bc7fe2301… |
| llm_5k_gemini-2.5-flash_fp_run1 | gemini-2.5-flash | fp | skills/llm_instructions_5k.md | 5774 | resolved | 4621a8e54f5d… |
| llm_5k_gemini-2.5-flash_nfp_run1 | gemini-2.5-flash | nfp | skills/llm_instructions_5k.md | 5774 | resolved | c7581cd57b47… |
| llm_5k_gemini-3.5-flash_fp_run1 | gemini-3.5-flash | fp | skills/llm_instructions_5k.md | 5774 | resolved | 21b20cfb9a4b… |
| llm_5k_gemini-3.5-flash_nfp_run1 | gemini-3.5-flash | nfp | skills/llm_instructions_5k.md | 5774 | resolved | d2a8898ef47f… |
| llm_5k_gpt-4o-2024-11-20_fp_run1 | gpt-4o-2024-11-20 | fp | skills/llm_instructions_5k.md | 5774 | resolved | 11f8497cbbf0… |
| llm_5k_gpt-4o-2024-11-20_nfp_run1 | gpt-4o-2024-11-20 | nfp | skills/llm_instructions_5k.md | 5774 | resolved | 8992dd04c16e… |
| llm_5k_gpt-5.4_fp_run1 | gpt-5.4 | fp | skills/llm_instructions_5k.md | 5774 | resolved | 0e10c1c7e89a… |
| llm_5k_gpt-5.4_nfp_run1 | gpt-5.4 | nfp | skills/llm_instructions_5k.md | 5774 | resolved | a50b5f188ead… |
| llm_5k_gpt-5.6-luna_fp_run1 | gpt-5.6-luna | fp | skills/llm_instructions_5k.md | 5774 | resolved | 63f3cdfdba61… |
| llm_5k_gpt-5.6-luna_nfp_run1 | gpt-5.6-luna | nfp | skills/llm_instructions_5k.md | 5774 | resolved | 924ff1b25dd5… |

| Condition | Model | Type | Full artifact | Chars | Lineage | Ledger SHA-256 |
|---|---|---|---|---|---|---|
| llm_5k_us.meta.llama3-1-8b-instruct-v1:0_fp_run1 | us.meta.llama3-1-8b-instruct-v1:0 | fp | skills/llm_instructions_5k.md | 5774 | resolved | 88bcf25bdf32… |
| llm_5k_us.meta.llama3-1-8b-instruct-v1:0_nfp_run1 | us.meta.llama3-1-8b-instruct-v1:0 | nfp | skills/llm_instructions_5k.md | 5774 | resolved | 03cbe502a2ab… |
| llm_5k_us.meta.llama3-3-70b-instruct-v1:0_fp_run1 | us.meta.llama3-3-70b-instruct-v1:0 | fp | skills/llm_instructions_5k.md | 5774 | resolved | 6199e221d9c7… |
| llm_5k_us.meta.llama3-3-70b-instruct-v1:0_nfp_run1 | us.meta.llama3-3-70b-instruct-v1:0 | nfp | skills/llm_instructions_5k.md | 5774 | resolved | 7b8eaded64df… |
| llm_natural_gemini-2.5-flash_fp_run1 | gemini-2.5-flash | fp | skills/llm_instructions_natural.md | 14533 | resolved | 80f11553b880… |
| llm_natural_gemini-2.5-flash_nfp_run1 | gemini-2.5-flash | nfp | skills/llm_instructions_natural.md | 14533 | resolved | f5bc34131f25… |
| llm_natural_gemini-3.5-flash_fp_run1 | gemini-3.5-flash | fp | skills/llm_instructions_natural.md | 14533 | resolved | ed0d1e4f2325… |
| llm_natural_gemini-3.5-flash_nfp_run1 | gemini-3.5-flash | nfp | skills/llm_instructions_natural.md | 14533 | resolved | 7e27259fec34… |
| llm_natural_gpt-4o-2024-11-20_fp_run1 | gpt-4o-2024-11-20 | fp | skills/llm_instructions_natural.md | 14533 | resolved | 8e55ce64f43f… |
| llm_natural_gpt-4o-2024-11-20_nfp_run1 | gpt-4o-2024-11-20 | nfp | skills/llm_instructions_natural.md | 14533 | resolved | 996891cc2804… |
| llm_natural_gpt-5.4_fp_run1 | gpt-5.4 | fp | skills/llm_instructions_natural.md | 14533 | resolved | e3143cae13f6… |
| llm_natural_gpt-5.4_nfp_run1 | gpt-5.4 | nfp | skills/llm_instructions_natural.md | 14533 | resolved | 9ef495ff7465… |
| llm_natural_gpt-5.6-luna_fp_run1 | gpt-5.6-luna | fp | skills/llm_instructions_natural.md | 14533 | resolved | df8ddc9b3101… |
| llm_natural_gpt-5.6-luna_nfp_run1 | gpt-5.6-luna | nfp | skills/llm_instructions_natural.md | 14533 | resolved | cd240628a0fa… |
| llm_natural_us.meta.llama3-1-8b-instruct-v1:0_fp_run1 | us.meta.llama3-1-8b-instruct-v1:0 | fp | skills/llm_instructions_natural.md | 14533 | resolved | 55af452329b9… |
| llm_natural_us.meta.llama3-1-8b-instruct-v1:0_nfp_run1 | us.meta.llama3-1-8b-instruct-v1:0 | nfp | skills/llm_instructions_natural.md | 14533 | resolved | 80ab5a7ec95a… |
| llm_natural_us.meta.llama3-3-70b-instruct-v1:0_fp_run1 | us.meta.llama3-3-70b-instruct-v1:0 | fp | skills/llm_instructions_natural.md | 14533 | resolved | 14354dcc85cf… |
| llm_natural_us.meta.llama3-3-70b-instruct-v1:0_nfp_run1 | us.meta.llama3-3-70b-instruct-v1:0 | nfp | skills/llm_instructions_natural.md | 14533 | resolved | e90683c110ba… |
| luna_fewshot_gpt-5.6-luna_fp_run1 | gpt-5.6-luna | fp | skills/fewshot_gpt54.md | 5927 | resolved | a703467e4108… |
| luna_fewshot_gpt-5.6-luna_nfp_run1 | gpt-5.6-luna | nfp | skills/fewshot_gpt54.md | 5927 | resolved | cf835e1241e6… |
| luna_gpt4o_skill_gpt-5.6-luna_fp_run1 | gpt-5.6-luna | fp | results/skillopt_v2_gpt4o/best_skill.md | 4933 | resolved | 70dfe4854f85… |
| luna_gpt4o_skill_gpt-5.6-luna_nfp_run1 | gpt-5.6-luna | nfp | results/skillopt_v2_gpt4o/best_skill.md | 4933 | resolved | 3a2021ba038a… |
| luna_opt_skill_gpt-4o-2024-11-20_nfp_run1 | gpt-4o-2024-11-20 | nfp | results/skillopt_v2_gpt4o_luna_opt/best_skill.md | 7051 | resolved | 5ff4c4ead916… |
| port_gemini_skill_gpt-4o-2024-11-20_fp_run1 | gpt-4o-2024-11-20 | fp | results/skillopt_v2_gemini_flash/best_skill.md | 4806 | resolved | f755d2daea5c… |
| port_gemini_skill_gpt-4o-2024-11-20_nfp_run1 | gpt-4o-2024-11-20 | nfp | results/skillopt_v2_gemini_flash/best_skill.md | 4806 | resolved | 9bf1556b5a5b… |
| port_gpt4o_skill_gemini-2.5-flash_fp_run1 | gemini-2.5-flash | fp | results/skillopt_v2_gpt4o/best_skill.md | 4933 | resolved | 00c2f9e32413… |
| port_gpt4o_skill_gemini-2.5-flash_nfp_run1 | gemini-2.5-flash | nfp | results/skillopt_v2_gpt4o/best_skill.md | 4933 | resolved | c4a94c6c4668… |
| port_gpt4o_skill_gemini-3.5-flash_fp_run1 | gemini-3.5-flash | fp | results/skillopt_v2_gpt4o/best_skill.md | 4933 | resolved | abdb11c1e3db… |
| port_gpt4o_skill_gemini-3.5-flash_nfp_run1 | gemini-3.5-flash | nfp | results/skillopt_v2_gpt4o/best_skill.md | 4933 | resolved | 4779ae53c6b0… |
| port_gpt4o_skill_us.meta.llama3-1-8b-instruct-v1:0_fp_run1 | us.meta.llama3-1-8b-instruct-v1:0 | fp | results/skillopt_v2_gpt4o/best_skill.md | 4933 | resolved | 633c177fe2c7… |
| port_gpt4o_skill_us.meta.llama3-1-8b-instruct-v1:0_nfp_run1 | us.meta.llama3-1-8b-instruct-v1:0 | nfp | results/skillopt_v2_gpt4o/best_skill.md | 4933 | resolved | b591a138801e… |
| port_gpt4o_skill_us.meta.llama3-3-70b-instruct-v1:0_fp_run1 | us.meta.llama3-3-70b-instruct-v1:0 | fp | results/skillopt_v2_gpt4o/best_skill.md | 4933 | resolved | f66cf14fe0ee… |
| port_gpt4o_skill_us.meta.llama3-3-70b-instruct-v1:0_nfp_run1 | us.meta.llama3-3-70b-instruct-v1:0 | nfp | results/skillopt_v2_gpt4o/best_skill.md | 4933 | resolved | 801c3e392462… |
| port_gpt54_skill_gemini-2.5-flash_fp_run1 | gemini-2.5-flash | fp | results/skillopt_v2_gpt54/best_skill.md | 5352 | resolved | a7615ae66c76… |
| port_gpt54_skill_gemini-2.5-flash_nfp_run1 | gemini-2.5-flash | nfp | results/skillopt_v2_gpt54/best_skill.md | 5352 | resolved | f97c1c8d182e… |
| port_gpt54_skill_gpt-4o-2024-11-20_fp_run1 | gpt-4o-2024-11-20 | fp | results/skillopt_v2_gpt54/best_skill.md | 5352 | resolved | 8afceb3cb44e… |
| port_gpt54_skill_gpt-4o-2024-11-20_nfp_run1 | gpt-4o-2024-11-20 | nfp | results/skillopt_v2_gpt54/best_skill.md | 5352 | resolved | 4caf265a825b… |
| port_rep1_skill_gemini-3.5-flash_fp_run1 | gemini-3.5-flash | fp | results/skillopt_v2_gpt4o_replicate1/best_skill.md | 4414 | resolved | 41441569f8f5… |
| port_rep1_skill_gpt-5.4_fp_run1 | gpt-5.4 | fp | results/skillopt_v2_gpt4o_replicate1/best_skill.md | 4414 | resolved | fa25140e6b92… |
| port_rep1_skill_us.meta.llama3-3-70b-instruct-v1:0_fp_run1 | us.meta.llama3-3-70b-instruct-v1:0 | fp | results/skillopt_v2_gpt4o_replicate1/best_skill.md | 4414 | resolved | 595a238dd739… |
| port_rep2_skill_gemini-3.5-flash_fp_run1 | gemini-3.5-flash | fp | results/skillopt_v2_gpt4o_replicate2/best_skill.md | 5416 | resolved | c6305b51afa7… |
| port_rep2_skill_gpt-5.4_fp_run1 | gpt-5.4 | fp | results/skillopt_v2_gpt4o_replicate2/best_skill.md | 5416 | resolved | 55537ab946c4… |
| port_rep2_skill_us.meta.llama3-3-70b-instruct-v1:0_fp_run1 | us.meta.llama3-3-70b-instruct-v1:0 | fp | results/skillopt_v2_gpt4o_replicate2/best_skill.md | 5416 | resolved | 752a4a546562… |
| skillopt_v0001_gpt-4o-2024-11-20_fp_run1 | gpt-4o-2024-11-20 | fp | results/skillopt_run/skills/skill_v0001.md | 3779 | resolved | d50236cd0021… |
| skillopt_v0001_gpt-4o-2024-11-20_nfp_run1 | gpt-4o-2024-11-20 | nfp | results/skillopt_run/skills/skill_v0001.md | 3779 | resolved | cc8de53d6005… |

| Condition | Model | Type | Full artifact | Chars | Lineage | Ledger SHA-256 |
|---|---|---|---|---|---|---|
| skillopt_v2_best_llama-3.1-8b_nfp_run1 | llama-3.1-8b | nfp | target-specific SkillOpt run directory | – | resolved by named run label; source differs by target model | 08311f711919… |
| skillopt_v2_gemini_best_gemini-2.5-flash_nfp_run1 | gemini-2.5-flash | nfp | target-specific SkillOpt run directory | – | resolved by named run label; source differs by target model | 0edb1ec3ba1f… |
| skillopt_v2_gpt4o_best_gpt-4o-2024-11-20_fp_run1 | gpt-4o-2024-11-20 | fp | target-specific SkillOpt run directory | – | resolved by named run label; source differs by target model | 6bc57dd2595e… |
| skillopt_v2_gpt4o_best_gpt-4o-2024-11-20_nfp_run1 | gpt-4o-2024-11-20 | nfp | target-specific SkillOpt run directory | – | resolved by named run label; source differs by target model | a5746f04348a… |
| skillopt_v2_gpt54_best_gpt-5.4_nfp_run1 | gpt-5.4 | nfp | target-specific SkillOpt run directory | – | resolved by named run label; source differs by target model | a9a304b9f14e… |

### S6. Complete cross-model results

Every eligible 585-question FP ledger was restricted to the same 410-question FP test set. Every eligible 150-question NFP ledger was restricted to the 120 controls not used for selection. Missing records, inference errors and invalid judge scores were counted as failures. Available-case estimates excluded these observations.

#### Table S10. All 123 common-population secondary results

| Family | Intervention | Model | Outcome | N | Success | Conservative % (95% CI) | Available % | Missing | SHA-256 |
|---|---|---|---|---|---|---|---|---|---|
| Baseline capability | No added skill | gpt-5.4 | Correction rate | 410 | 221 | 53.9 (49.1–58.7) | 53.9 | 0 | 1607587ca886… |
| Baseline capability | No added skill | gpt-5.4 | Negative-control specificity | 120 | 108 | 90.0 (83.3–94.2) | 90.0 | 0 | 327cb2d2e13e… |
| Baseline capability | No added skill | gemini-2.5-flash | Correction rate | 410 | 0 | 0.0 (0.0–0.9) | 0.0 | 11 | 0262b9ba742f… |
| Baseline capability | No added skill | gemini-2.5-flash | Negative-control specificity | 120 | 75 | 62.5 (53.6–70.6) | 64.7 | 4 | b40f13ae62ad… |
| Baseline capability | No added skill | gemini-3.5-flash | Correction rate | 410 | 242 | 59.0 (54.2–63.7) | 59.0 | 0 | 66eb3a55b43d… |
| Baseline capability | No added skill | gemini-3.5-flash | Negative-control specificity | 120 | 103 | 85.8 (78.5–91.0) | 85.8 | 0 | cf897f61b579… |
| Baseline capability | No added skill | gpt-4o-2024-11-20 | Correction rate | 410 | 18 | 4.4 (2.8–6.8) | 4.4 | 0 | e2c970dea97d… |
| Baseline capability | No added skill | gpt-4o-2024-11-20 | Negative-control specificity | 120 | 92 | 76.7 (68.3–83.3) | 76.7 | 0 | 395fcd2d1612… |
| Baseline capability | No added skill | llama-3.1-8b | Negative-control specificity | 120 | 84 | 70.0 (61.3–77.5) | 70.0 | 0 | 157533b0702e… |
| Baseline capability | No added skill | gpt-5.6-luna | Correction rate | 410 | 293 | 71.5 (66.9–75.6) | 71.5 | 0 | bd3ed1f8a3c8… |
| Baseline capability | No added skill | gpt-5.6-luna | Negative-control specificity | 120 | 115 | 95.8 (90.6–98.2) | 95.8 | 0 | da8e906d8d5b… |
| Baseline capability | No added skill | us.meta.llama3-1-8b-instruct-v1:0 | Correction rate | 410 | 1 | 0.2 (0.0–1.4) | 0.2 | 0 | b536db4b269a… |
| Baseline capability | No added skill | us.meta.llama3-1-8b-instruct-v1:0 | Negative-control specificity | 120 | 94 | 78.3 (70.1–84.8) | 78.3 | 0 | 538481e2bfc0… |
| Baseline capability | No added skill | us.meta.llama3-3-70b-instruct-v1:0 | Correction rate | 410 | 11 | 2.7 (1.5–4.7) | 2.7 | 0 | ad6f74e0907a… |
| Baseline capability | No added skill | us.meta.llama3-3-70b-instruct-v1:0 | Negative-control specificity | 120 | 73 | 60.8 (51.9–69.1) | 60.8 | 0 | ce4664da2fd1… |
| Composite intervention | Skill plus examples | gemini-2.5-flash | Correction rate | 410 | 198 | 48.3 (43.5–53.1) | 49.9 | 13 | 9647edb05fe3… |
| Composite intervention | Skill plus examples | gemini-2.5-flash | Negative-control specificity | 120 | 54 | 45.0 (36.4–53.9) | 45.0 | 0 | 61272d3e77e0… |
| Composite intervention | Skill plus examples | gpt-4o-2024-11-20 | Correction rate | 410 | 359 | 87.6 (84.0–90.4) | 87.6 | 0 | 9f30cadb064e… |
| Composite intervention | Skill plus examples | gpt-4o-2024-11-20 | Negative-control specificity | 120 | 58 | 48.3 (39.6–57.2) | 48.3 | 0 | df4cb448acbe… |
| Compression | Compact learned skill | gemini-2.5-flash | Correction rate | 410 | 41 | 10.0 (7.5–13.3) | 10.6 | 23 | 43c94262632c… |
| Compression | Compact learned skill | gemini-2.5-flash | Negative-control specificity | 120 | 64 | 53.3 (44.4–62.0) | 54.2 | 2 | cfc8815a2e75… |
| Compression | Compact learned skill | gemini-3.5-flash | Correction rate | 410 | 369 | 90.0 (86.7–92.5) | 90.0 | 0 | f7718ee1e19d… |
| Compression | Compact learned skill | gemini-3.5-flash | Negative-control specificity | 120 | 103 | 85.8 (78.5–91.0) | 85.8 | 0 | 9374ef5d8111… |
| Compression | Compact learned skill | gpt-4o-2024-11-20 | Correction rate | 410 | 231 | 56.3 (51.5–61.1) | 56.3 | 0 | caabed18f0ef… |
| Compression | Compact learned skill | gpt-4o-2024-11-20 | Negative-control specificity | 120 | 95 | 79.2 (71.1–85.5) | 79.2 | 0 | d5f0eb03536e… |
| Compression | Compact learned skill | gpt-5.4 | Correction rate | 410 | 370 | 90.2 (87.0–92.8) | 90.2 | 0 | c42482443049… |
| Compression | Compact learned skill | gpt-5.4 | Negative-control specificity | 120 | 104 | 86.7 (79.4–91.6) | 86.7 | 0 | 6d30787b4251… |
| Compression | Compact learned skill | gpt-5.6-luna | Correction rate | 410 | 381 | 92.9 (90.0–95.0) | 92.9 | 0 | 8b7d8407c860… |
| Compression | Compact learned skill | gpt-5.6-luna | Negative-control specificity | 120 | 112 | 93.3 (87.4–96.6) | 93.3 | 0 | 120cde872f6c… |
| Compression | Compact learned skill | us.meta.llama3-1-8b-instruct-v1:0 | Correction rate | 410 | 28 | 6.8 (4.8–9.7) | 6.8 | 0 | 25f32442f893… |
| Compression | Compact learned skill | us.meta.llama3-1-8b-instruct-v1:0 | Negative-control specificity | 120 | 87 | 72.5 (63.9–79.7) | 72.5 | 0 | c6e1009ff4cf… |
| Compression | Compact learned skill | us.meta.llama3-3-70b-instruct-v1:0 | Correction rate | 410 | 220 | 53.7 (48.8–58.4) | 53.7 | 0 | b662b5080202… |
| Compression | Compact learned skill | us.meta.llama3-3-70b-instruct-v1:0 | Negative-control specificity | 120 | 71 | 59.2 (50.2–67.5) | 59.2 | 0 | f80225b9183a… |
| Prompt strategy | Size-matched few-shot examples | gemini-2.5-flash | Correction rate | 410 | 224 | 54.6 (49.8–59.4) | 55.7 | 8 | 1c59172f5c30… |
| Prompt strategy | Size-matched few-shot examples | gemini-2.5-flash | Negative-control specificity | 120 | 61 | 50.8 (42.0–59.6) | 50.8 | 0 | 937fa227254c… |
| Prompt strategy | Size-matched few-shot examples | gemini-3.5-flash | Correction rate | 410 | 363 | 88.5 (85.1–91.3) | 88.5 | 0 | 93a878ce53a8… |
| Prompt strategy | Size-matched few-shot examples | gemini-3.5-flash | Negative-control specificity | 120 | 68 | 56.7 (47.7–65.2) | 56.7 | 0 | 03613e4c53fe… |

| Family | Intervention | Model | Outcome | N | Success | Conservative % (95% CI) | Available % | Missing | SHA-256 |
|---|---|---|---|---|---|---|---|---|---|
| Prompt strategy | Size-matched few-shot examples | gpt-4o-2024-11-20 | Correction rate | 410 | 306 | 74.6 (70.2–78.6) | 74.6 | 0 | ea9fc5a5e8d7… |
| Prompt strategy | Size-matched few-shot examples | gpt-4o-2024-11-20 | Negative-control specificity | 120 | 78 | 65.0 (56.1–72.9) | 65.0 | 0 | 5d921f4e9498… |
| Prompt strategy | Size-matched few-shot examples | llama-3.1-8b | Correction rate | 410 | 220 | 53.7 (48.8–58.4) | 53.8 | 1 | 2038292e2c69… |
| Prompt strategy | Size-matched few-shot examples | llama-3.1-8b | Negative-control specificity | 120 | 89 | 74.2 (65.7–81.2) | 74.2 | 0 | c70fcbdef35f… |
| Prompt strategy | Size-matched few-shot examples | us.meta.llama3-1-8b-instruct-v1:0 | Correction rate | 410 | 183 | 44.6 (39.9–49.5) | 44.6 | 0 | 3d3cb44af552… |
| Prompt strategy | Size-matched few-shot examples | us.meta.llama3-1-8b-instruct-v1:0 | Negative-control specificity | 120 | 47 | 39.2 (30.9–48.1) | 39.2 | 0 | 9e083dde0987… |
| Prompt strategy | Size-matched few-shot examples | us.meta.llama3-3-70b-instruct-v1:0 | Correction rate | 410 | 268 | 65.4 (60.6–69.8) | 65.4 | 0 | 4e0f52ca326c… |
| Prompt strategy | Size-matched few-shot examples | us.meta.llama3-3-70b-instruct-v1:0 | Negative-control specificity | 120 | 48 | 40.0 (31.7–48.9) | 40.0 | 0 | 3a4c776fb4c6… |
| Prompt strategy | Size-matched few-shot examples | gpt-5.4 | Correction rate | 410 | 369 | 90.0 (86.7–92.5) | 90.0 | 0 | ca2905c43b6c… |
| Prompt strategy | Size-matched few-shot examples | gpt-5.4 | Negative-control specificity | 120 | 95 | 79.2 (71.1–85.5) | 79.2 | 0 | 7b90bd6fff7b… |
| Skill portability | Primary GPT-4o learned skill | gpt-5.4 | Correction rate | 410 | 383 | 93.4 (90.6–95.4) | 93.4 | 0 | 1462bfe32496… |
| Skill portability | Primary GPT-4o learned skill | gpt-5.4 | Negative-control specificity | 120 | 101 | 84.2 (76.6–89.6) | 84.2 | 0 | 0063d7074ea3… |
| Prompt strategy | Approximately 2k-character instruction | gemini-2.5-flash | Correction rate | 410 | 86 | 21.0 (17.3–25.2) | 21.9 | 18 | 8674067d7eaa… |
| Prompt strategy | Approximately 2k-character instruction | gemini-2.5-flash | Negative-control specificity | 120 | 68 | 56.7 (47.7–65.2) | 58.1 | 3 | 77a475dd7c1d… |
| Prompt strategy | Approximately 2k-character instruction | gemini-3.5-flash | Correction rate | 410 | 374 | 91.2 (88.1–93.6) | 91.2 | 0 | 7eb9d8369590… |
| Prompt strategy | Approximately 2k-character instruction | gemini-3.5-flash | Negative-control specificity | 120 | 93 | 77.5 (69.2–84.1) | 77.5 | 0 | 89f487bf20e6… |
| Prompt strategy | Approximately 2k-character instruction | gpt-4o-2024-11-20 | Correction rate | 410 | 280 | 68.3 (63.6–72.6) | 68.3 | 0 | 6b67d1add93d… |
| Prompt strategy | Approximately 2k-character instruction | gpt-4o-2024-11-20 | Negative-control specificity | 120 | 91 | 75.8 (67.5–82.6) | 75.8 | 0 | 56de14fead06… |
| Prompt strategy | Approximately 2k-character instruction | gpt-5.4 | Correction rate | 410 | 351 | 85.6 (81.9–88.7) | 85.6 | 0 | b971b840b78d… |
| Prompt strategy | Approximately 2k-character instruction | gpt-5.4 | Negative-control specificity | 120 | 104 | 86.7 (79.4–91.6) | 86.7 | 0 | 96d323111c57… |
| Prompt strategy | Approximately 2k-character instruction | gpt-5.6-luna | Correction rate | 410 | 371 | 90.5 (87.3–93.0) | 90.5 | 0 | 54f4a55d0463… |
| Prompt strategy | Approximately 2k-character instruction | gpt-5.6-luna | Negative-control specificity | 120 | 103 | 85.8 (78.5–91.0) | 85.8 | 0 | e7cf2b668cab… |
| Prompt strategy | Approximately 2k-character instruction | us.meta.llama3-1-8b-instruct-v1:0 | Correction rate | 410 | 167 | 40.7 (36.1–45.6) | 40.7 | 0 | 346e3e595063… |
| Prompt strategy | Approximately 2k-character instruction | us.meta.llama3-1-8b-instruct-v1:0 | Negative-control specificity | 120 | 50 | 41.7 (33.2–50.6) | 41.7 | 0 | dba810853c65… |
| Prompt strategy | Approximately 2k-character instruction | us.meta.llama3-3-70b-instruct-v1:0 | Correction rate | 410 | 173 | 42.2 (37.5–47.0) | 42.2 | 0 | da21951a68ab… |
| Prompt strategy | Approximately 2k-character instruction | us.meta.llama3-3-70b-instruct-v1:0 | Negative-control specificity | 120 | 77 | 64.2 (55.3–72.2) | 64.2 | 0 | c71bc7fe2301… |
| Prompt strategy | Approximately 5k-character instruction | gemini-2.5-flash | Correction rate | 410 | 74 | 18.0 (14.6–22.1) | 19.0 | 21 | 4621a8e54f5d… |
| Prompt strategy | Approximately 5k-character instruction | gemini-2.5-flash | Negative-control specificity | 120 | 75 | 62.5 (53.6–70.6) | 63.0 | 1 | c7581cd57b47… |
| Prompt strategy | Approximately 5k-character instruction | gemini-3.5-flash | Correction rate | 410 | 375 | 91.5 (88.4–93.8) | 91.5 | 0 | 21b20cfb9a4b… |
| Prompt strategy | Approximately 5k-character instruction | gemini-3.5-flash | Negative-control specificity | 120 | 97 | 80.8 (72.9–86.9) | 80.8 | 0 | d2a8898ef47f… |
| Prompt strategy | Approximately 5k-character instruction | gpt-4o-2024-11-20 | Correction rate | 410 | 300 | 73.2 (68.7–77.2) | 73.2 | 0 | 11f8497cbbf0… |
| Prompt strategy | Approximately 5k-character instruction | gpt-4o-2024-11-20 | Negative-control specificity | 120 | 81 | 67.5 (58.7–75.2) | 67.5 | 0 | 8992dd04c16e… |
| Prompt strategy | Approximately 5k-character instruction | gpt-5.4 | Correction rate | 410 | 379 | 92.4 (89.5–94.6) | 92.4 | 0 | 0e10c1c7e89a… |
| Prompt strategy | Approximately 5k-character instruction | gpt-5.4 | Negative-control specificity | 120 | 107 | 89.2 (82.3–93.6) | 89.2 | 0 | a50b5f188ead… |
| Prompt strategy | Approximately 5k-character instruction | gpt-5.6-luna | Correction rate | 410 | 372 | 90.7 (87.5–93.2) | 90.7 | 0 | 63f3cdfdba61… |
| Prompt strategy | Approximately 5k-character instruction | gpt-5.6-luna | Negative-control specificity | 120 | 105 | 87.5 (80.4–92.3) | 87.5 | 0 | 924ff1b25dd5… |
| Prompt strategy | Approximately 5k-character instruction | us.meta.llama3-1-8b-instruct-v1:0 | Correction rate | 410 | 63 | 15.4 (12.2–19.2) | 15.4 | 0 | 88bcf25bdf32… |
| Prompt strategy | Approximately 5k-character instruction | us.meta.llama3-1-8b-instruct-v1:0 | Negative-control specificity | 120 | 58 | 48.3 (39.6–57.2) | 48.3 | 0 | 03cbe502a2ab… |

| Family | Intervention | Model | Outcome | N | Success | Conservative % (95% CI) | Available % | Missing | SHA-256 |
|---|---|---|---|---|---|---|---|---|---|
| Prompt strategy | Approximately 5k-character instruction | us.meta.llama3-3-70b-instruct-v1:0 | Correction rate | 410 | 166 | 40.5 (35.8–45.3) | 40.5 | 0 | 6199e221d9c7… |
| Prompt strategy | Approximately 5k-character instruction | us.meta.llama3-3-70b-instruct-v1:0 | Negative-control specificity | 120 | 75 | 62.5 (53.6–70.6) | 62.5 | 0 | 7b8eaded64df… |
| Prompt strategy | Natural-language instruction | gemini-2.5-flash | Correction rate | 410 | 23 | 5.6 (3.8–8.3) | 6.0 | 25 | 80f11553b880… |
| Prompt strategy | Natural-language instruction | gemini-2.5-flash | Negative-control specificity | 120 | 74 | 61.7 (52.7–69.9) | 62.7 | 2 | f5bc34131f25… |
| Prompt strategy | Natural-language instruction | gemini-3.5-flash | Correction rate | 410 | 375 | 91.5 (88.4–93.8) | 91.5 | 0 | ed0d1e4f2325… |
| Prompt strategy | Natural-language instruction | gemini-3.5-flash | Negative-control specificity | 120 | 95 | 79.2 (71.1–85.5) | 79.2 | 0 | 7e27259fec34… |
| Prompt strategy | Natural-language instruction | gpt-4o-2024-11-20 | Correction rate | 410 | 251 | 61.2 (56.4–65.8) | 61.2 | 0 | 8e55ce64f43f… |
| Prompt strategy | Natural-language instruction | gpt-4o-2024-11-20 | Negative-control specificity | 120 | 93 | 77.5 (69.2–84.1) | 77.5 | 0 | 996891cc2804… |
| Prompt strategy | Natural-language instruction | gpt-5.4 | Correction rate | 410 | 377 | 92.0 (88.9–94.2) | 92.0 | 0 | e3143cae13f6… |
| Prompt strategy | Natural-language instruction | gpt-5.4 | Negative-control specificity | 120 | 106 | 88.3 (81.4–92.9) | 88.3 | 0 | 9ef495ff7465… |
| Prompt strategy | Natural-language instruction | gpt-5.6-luna | Correction rate | 410 | 366 | 89.3 (85.9–91.9) | 89.3 | 0 | df8ddc9b3101… |
| Prompt strategy | Natural-language instruction | gpt-5.6-luna | Negative-control specificity | 120 | 111 | 92.5 (86.4–96.0) | 92.5 | 0 | cd240628a0fa… |
| Prompt strategy | Natural-language instruction | us.meta.llama3-1-8b-instruct-v1:0 | Correction rate | 410 | 37 | 9.0 (6.6–12.2) | 9.0 | 0 | 55af452329b9… |
| Prompt strategy | Natural-language instruction | us.meta.llama3-1-8b-instruct-v1:0 | Negative-control specificity | 120 | 66 | 55.0 (46.1–63.6) | 55.0 | 0 | 80ab5a7ec95a… |
| Prompt strategy | Natural-language instruction | us.meta.llama3-3-70b-instruct-v1:0 | Correction rate | 410 | 107 | 26.1 (22.1–30.6) | 26.1 | 0 | 14354dcc85cf… |
| Prompt strategy | Natural-language instruction | us.meta.llama3-3-70b-instruct-v1:0 | Negative-control specificity | 120 | 74 | 61.7 (52.7–69.9) | 61.7 | 0 | e90683c110ba… |
| Prompt strategy | Size-matched few-shot examples | gpt-5.6-luna | Correction rate | 410 | 382 | 93.2 (90.3–95.2) | 93.2 | 0 | a703467e4108… |
| Prompt strategy | Size-matched few-shot examples | gpt-5.6-luna | Negative-control specificity | 120 | 96 | 80.0 (72.0–86.2) | 80.0 | 0 | cf835e1241e6… |
| Skill portability | Primary GPT-4o learned skill | gpt-5.6-luna | Correction rate | 410 | 396 | 96.6 (94.4–98.0) | 96.6 | 0 | 70dfe4854f85… |
| Skill portability | Primary GPT-4o learned skill | gpt-5.6-luna | Negative-control specificity | 120 | 95 | 79.2 (71.1–85.5) | 79.2 | 0 | 3a2021ba038a… |
| Skill portability | Luna-optimised GPT-4o skill | gpt-4o-2024-11-20 | Negative-control specificity | 120 | 92 | 76.7 (68.3–83.3) | 76.7 | 0 | 5ff4c4ead916… |
| Skill portability | Gemini learned skill | gpt-4o-2024-11-20 | Correction rate | 410 | 336 | 82.0 (77.9–85.4) | 82.0 | 0 | f755d2daea5c… |
| Skill portability | Gemini learned skill | gpt-4o-2024-11-20 | Negative-control specificity | 120 | 79 | 65.8 (57.0–73.7) | 65.8 | 0 | 9bf1556b5a5b… |
| Skill portability | Primary GPT-4o learned skill | gemini-2.5-flash | Correction rate | 410 | 26 | 6.3 (4.4–9.1) | 6.7 | 19 | 00c2f9e32413… |
| Skill portability | Primary GPT-4o learned skill | gemini-2.5-flash | Negative-control specificity | 120 | 69 | 57.5 (48.6–66.0) | 58.5 | 2 | c4a94c6c4668… |
| Skill portability | Primary GPT-4o learned skill | gemini-3.5-flash | Correction rate | 410 | 380 | 92.7 (89.7–94.8) | 92.7 | 0 | abdb11c1e3db… |
| Skill portability | Primary GPT-4o learned skill | gemini-3.5-flash | Negative-control specificity | 120 | 71 | 59.2 (50.2–67.5) | 59.2 | 0 | 4779ae53c6b0… |
| Skill portability | Primary GPT-4o learned skill | us.meta.llama3-1-8b-instruct-v1:0 | Correction rate | 410 | 67 | 16.3 (13.1–20.2) | 16.3 | 0 | 633c177fe2c7… |
| Skill portability | Primary GPT-4o learned skill | us.meta.llama3-1-8b-instruct-v1:0 | Negative-control specificity | 120 | 72 | 60.0 (51.1–68.3) | 60.0 | 0 | b591a138801e… |
| Skill portability | Primary GPT-4o learned skill | us.meta.llama3-3-70b-instruct-v1:0 | Correction rate | 410 | 226 | 55.1 (50.3–59.9) | 55.1 | 0 | f66cf14fe0ee… |
| Skill portability | Primary GPT-4o learned skill | us.meta.llama3-3-70b-instruct-v1:0 | Negative-control specificity | 120 | 60 | 50.0 (41.2–58.8) | 50.0 | 0 | 801c3e392462… |
| Skill portability | GPT-5.4 learned skill | gemini-2.5-flash | Correction rate | 410 | 12 | 2.9 (1.7–5.0) | 3.1 | 22 | a7615ae66c76… |
| Skill portability | GPT-5.4 learned skill | gemini-2.5-flash | Negative-control specificity | 120 | 72 | 60.0 (51.1–68.3) | 60.0 | 0 | f97c1c8d182e… |
| Skill portability | GPT-5.4 learned skill | gpt-4o-2024-11-20 | Correction rate | 410 | 299 | 72.9 (68.4–77.0) | 72.9 | 0 | 8afceb3cb44e… |
| Skill portability | GPT-5.4 learned skill | gpt-4o-2024-11-20 | Negative-control specificity | 120 | 85 | 70.8 (62.2–78.2) | 70.8 | 0 | 4caf265a825b… |
| Skill portability | GPT-4o learned skill replicate 1 | gemini-3.5-flash | Correction rate | 410 | 395 | 96.3 (94.1–97.8) | 96.3 | 0 | 41441569f8f5… |
| Skill portability | GPT-4o learned skill replicate 1 | gpt-5.4 | Correction rate | 410 | 397 | 96.8 (94.7–98.1) | 97.1 | 1 | fa25140e6b92… |
| Skill portability | GPT-4o learned skill replicate 1 | us.meta.llama3-3-70b-instruct-v1:0 | Correction rate | 410 | 319 | 77.8 (73.5–81.6) | 77.8 | 0 | 595a238dd739… |
| Skill portability | GPT-4o learned skill replicate 2 | gemini-3.5-flash | Correction rate | 410 | 392 | 95.6 (93.2–97.2) | 95.6 | 0 | c6305b51afa7… |
| Skill portability | GPT-4o learned skill replicate 2 | gpt-5.4 | Correction rate | 410 | 399 | 97.3 (95.3–98.5) | 97.6 | 1 | 55537ab946c4… |
| Skill portability | GPT-4o learned skill replicate 2 | us.meta.llama3-3-70b-instruct-v1:0 | Correction rate | 410 | 250 | 61.0 (56.2–65.6) | 61.0 | 0 | 752a4a546562… |

| Family | Intervention | Model | Outcome | N | Success | Conservative % (95% CI) | Available % | Missing | SHA-256 |
|---|---|---|---|---|---|---|---|---|---|
| Legacy optimisation | Early SkillOpt candidate | gpt-4o-2024-11-20 | Correction rate | 410 | 260 | 63.4 (58.6–67.9) | 63.4 | 0 | d50236cd0021… |
| Legacy optimisation | Early SkillOpt candidate | gpt-4o-2024-11-20 | Negative-control specificity | 120 | 67 | 55.8 (46.9–64.4) | 55.8 | 0 | cc8de53d6005… |
| Skill evaluation | Named SkillOpt output | llama-3.1-8b | Negative-control specificity | 120 | 49 | 40.8 (32.5–49.8) | 40.8 | 0 | 08311f711919… |
| Skill evaluation | Named SkillOpt output | gemini-2.5-flash | Negative-control specificity | 120 | 72 | 60.0 (51.1–68.3) | 60.0 | 0 | 0edb1ec3ba1f… |
| Skill evaluation | Named SkillOpt output | gpt-4o-2024-11-20 | Correction rate | 410 | 382 | 93.2 (90.3–95.2) | 93.2 | 0 | 6bc57dd2595e… |
| Skill evaluation | Named SkillOpt output | gpt-4o-2024-11-20 | Negative-control specificity | 120 | 84 | 70.0 (61.3–77.5) | 70.0 | 0 | a5746f04348a… |
| Skill evaluation | Named SkillOpt output | gpt-5.4 | Negative-control specificity | 120 | 100 | 83.3 (75.7–88.9) | 83.3 | 0 | a9a304b9f14e… |

#### Table S11. Core comparison: baseline, few-shot, full SkillOpt and compact SkillOpt

| Model | Intervention | FP correction % (95% CI) | NFP accuracy % (95% CI) | Missing FP / NFP |
|---|---|---|---|---|
| Llama 3.1 8B | No added skill | 0.2 (0.0–1.4) | 78.3 (70.1–84.8) | 0 / 0 |
| Llama 3.1 8B | Size-matched few-shot examples | 44.6 (39.9–49.5) | 39.2 (30.9–48.1) | 0 / 0 |
| Llama 3.1 8B | Primary GPT-4o learned skill | 16.3 (13.1–20.2) | 60.0 (51.1–68.3) | 0 / 0 |
| Llama 3.1 8B | Compact learned skill | 6.8 (4.8–9.7) | 72.5 (63.9–79.7) | 0 / 0 |
| Gemini 2.5 Flash | No added skill | 0.0 (0.0–0.9) | 62.5 (53.6–70.6) | 11 / 4 |
| Gemini 2.5 Flash | Size-matched few-shot examples | 54.6 (49.8–59.4) | 50.8 (42.0–59.6) | 8 / 0 |
| Gemini 2.5 Flash | Primary GPT-4o learned skill | 6.3 (4.4–9.1) | 57.5 (48.6–66.0) | 19 / 2 |
| Gemini 2.5 Flash | Compact learned skill | 10.0 (7.5–13.3) | 53.3 (44.4–62.0) | 23 / 2 |
| Llama 3.3 70B | No added skill | 2.7 (1.5–4.7) | 60.8 (51.9–69.1) | 0 / 0 |
| Llama 3.3 70B | Size-matched few-shot examples | 65.4 (60.6–69.8) | 40.0 (31.7–48.9) | 0 / 0 |
| Llama 3.3 70B | Primary GPT-4o learned skill | 55.1 (50.3–59.9) | 50.0 (41.2–58.8) | 0 / 0 |
| Llama 3.3 70B | Compact learned skill | 53.7 (48.8–58.4) | 59.2 (50.2–67.5) | 0 / 0 |
| GPT-4o | No added skill | 4.4 (2.8–6.8) | 76.7 (68.3–83.3) | 0 / 0 |
| GPT-4o | Size-matched few-shot examples | 74.6 (70.2–78.6) | 65.0 (56.1–72.9) | 0 / 0 |
| GPT-4o | Compact learned skill | 56.3 (51.5–61.1) | 79.2 (71.1–85.5) | 0 / 0 |
| GPT-5.4 | No added skill | 53.9 (49.1–58.7) | 90.0 (83.3–94.2) | 0 / 0 |
| GPT-5.4 | Size-matched few-shot examples | 90.0 (86.7–92.5) | 79.2 (71.1–85.5) | 0 / 0 |
| GPT-5.4 | Primary GPT-4o learned skill | 93.4 (90.6–95.4) | 84.2 (76.6–89.6) | 0 / 0 |
| GPT-5.4 | Compact learned skill | 90.2 (87.0–92.8) | 86.7 (79.4–91.6) | 0 / 0 |
| Gemini 3.5 Flash | No added skill | 59.0 (54.2–63.7) | 85.8 (78.5–91.0) | 0 / 0 |
| Gemini 3.5 Flash | Size-matched few-shot examples | 88.5 (85.1–91.3) | 56.7 (47.7–65.2) | 0 / 0 |
| Gemini 3.5 Flash | Primary GPT-4o learned skill | 92.7 (89.7–94.8) | 59.2 (50.2–67.5) | 0 / 0 |
| Gemini 3.5 Flash | Compact learned skill | 90.0 (86.7–92.5) | 85.8 (78.5–91.0) | 0 / 0 |
| GPT-5.6 Luna | No added skill | 71.5 (66.9–75.6) | 95.8 (90.6–98.2) | 0 / 0 |
| GPT-5.6 Luna | Size-matched few-shot examples | 93.2 (90.3–95.2) | 80.0 (72.0–86.2) | 0 / 0 |
| GPT-5.6 Luna | Primary GPT-4o learned skill | 96.6 (94.4–98.0) | 79.2 (71.1–85.5) | 0 / 0 |
| GPT-5.6 Luna | Compact learned skill | 92.9 (90.0–95.0) | 93.3 (87.4–96.6) | 0 / 0 |

#### Table S12. Rule-prompt length and few-shot comparison

| Model | Intervention | FP correction % (95% CI) | NFP accuracy % (95% CI) | Missing FP / NFP |
|---|---|---|---|---|
| Llama 3.1 8B | Natural-language instruction | 9.0 (6.6–12.2) | 55.0 (46.1–63.6) | 0 / 0 |
| Llama 3.1 8B | Approximately 5k-character instruction | 15.4 (12.2–19.2) | 48.3 (39.6–57.2) | 0 / 0 |
| Llama 3.1 8B | Approximately 2k-character instruction | 40.7 (36.1–45.6) | 41.7 (33.2–50.6) | 0 / 0 |
| Llama 3.1 8B | Size-matched few-shot examples | 44.6 (39.9–49.5) | 39.2 (30.9–48.1) | 0 / 0 |
| Gemini 2.5 Flash | Natural-language instruction | 5.6 (3.8–8.3) | 61.7 (52.7–69.9) | 25 / 2 |
| Gemini 2.5 Flash | Approximately 5k-character instruction | 18.0 (14.6–22.1) | 62.5 (53.6–70.6) | 21 / 1 |
| Gemini 2.5 Flash | Approximately 2k-character instruction | 21.0 (17.3–25.2) | 56.7 (47.7–65.2) | 18 / 3 |
| Gemini 2.5 Flash | Size-matched few-shot examples | 54.6 (49.8–59.4) | 50.8 (42.0–59.6) | 8 / 0 |
| Llama 3.3 70B | Natural-language instruction | 26.1 (22.1–30.6) | 61.7 (52.7–69.9) | 0 / 0 |
| Llama 3.3 70B | Approximately 5k-character instruction | 40.5 (35.8–45.3) | 62.5 (53.6–70.6) | 0 / 0 |
| Llama 3.3 70B | Approximately 2k-character instruction | 42.2 (37.5–47.0) | 64.2 (55.3–72.2) | 0 / 0 |
| Llama 3.3 70B | Size-matched few-shot examples | 65.4 (60.6–69.8) | 40.0 (31.7–48.9) | 0 / 0 |
| GPT-4o | Natural-language instruction | 61.2 (56.4–65.8) | 77.5 (69.2–84.1) | 0 / 0 |
| GPT-4o | Approximately 5k-character instruction | 73.2 (68.7–77.2) | 67.5 (58.7–75.2) | 0 / 0 |
| GPT-4o | Approximately 2k-character instruction | 68.3 (63.6–72.6) | 75.8 (67.5–82.6) | 0 / 0 |
| GPT-4o | Size-matched few-shot examples | 74.6 (70.2–78.6) | 65.0 (56.1–72.9) | 0 / 0 |
| GPT-5.4 | Natural-language instruction | 92.0 (88.9–94.2) | 88.3 (81.4–92.9) | 0 / 0 |
| GPT-5.4 | Approximately 5k-character instruction | 92.4 (89.5–94.6) | 89.2 (82.3–93.6) | 0 / 0 |
| GPT-5.4 | Approximately 2k-character instruction | 85.6 (81.9–88.7) | 86.7 (79.4–91.6) | 0 / 0 |

| Model | Intervention | FP correction % (95% CI) | NFP accuracy % (95% CI) | Missing FP / NFP |
|---|---|---|---|---|
| GPT-5.4 | Size-matched few-shot examples | 90.0 (86.7–92.5) | 79.2 (71.1–85.5) | 0 / 0 |
| Gemini 3.5 Flash | Natural-language instruction | 91.5 (88.4–93.8) | 79.2 (71.1–85.5) | 0 / 0 |
| Gemini 3.5 Flash | Approximately 5k-character instruction | 91.5 (88.4–93.8) | 80.8 (72.9–86.9) | 0 / 0 |
| Gemini 3.5 Flash | Approximately 2k-character instruction | 91.2 (88.1–93.6) | 77.5 (69.2–84.1) | 0 / 0 |
| Gemini 3.5 Flash | Size-matched few-shot examples | 88.5 (85.1–91.3) | 56.7 (47.7–65.2) | 0 / 0 |
| GPT-5.6 Luna | Natural-language instruction | 89.3 (85.9–91.9) | 92.5 (86.4–96.0) | 0 / 0 |
| GPT-5.6 Luna | Approximately 5k-character instruction | 90.7 (87.5–93.2) | 87.5 (80.4–92.3) | 0 / 0 |
| GPT-5.6 Luna | Approximately 2k-character instruction | 90.5 (87.3–93.0) | 85.8 (78.5–91.0) | 0 / 0 |
| GPT-5.6 Luna | Size-matched few-shot examples | 93.2 (90.3–95.2) | 80.0 (72.0–86.2) | 0 / 0 |

##### Table S13. Composite skill-plus-example results

| Model | Outcome | Success / N | % (95% CI) | Missing |
|---|---|---|---|---|
| gemini-2.5-flash | Correction rate | 198 / 410 | 48.3 (43.5–53.1) | 13 |
| gemini-2.5-flash | Negative-control specificity | 54 / 120 | 45.0 (36.4–53.9) | 0 |
| gpt-4o-2024-11-20 | Correction rate | 359 / 410 | 87.6 (84.0–90.4) | 0 |
| gpt-4o-2024-11-20 | Negative-control specificity | 58 / 120 | 48.3 (39.6–57.2) | 0 |

#### S7. Thematic analysis and skill portability

Eight features were specified before comparing transfer results. A feature counted as present only when explicitly stated in the skill: premise checking; correction before advice; an explicit negate-then-replace template; conditional language; clinical determinants; oncology-team referral; avoidance of guidance that reinforced the FP; and supportive tone.

##### Table S14. Thematic coding of three independently learned GPT-4o skills

| Skill | Feature | Present | Matches | Characters |
|---|---|---|---|---|
| Primary skill | Premise audit | Yes | 14 | 4933 |
| Primary skill | Correction before advice | Yes | 3 | 4933 |
| Primary skill | Explicit negate-then-replace template | No | 0 | 4933 |
| Primary skill | Conditional framing | Yes | 15 | 4933 |
| Primary skill | Clinical determinants | Yes | 4 | 4933 |
| Primary skill | Oncology-team referral | Yes | 7 | 4933 |
| Primary skill | Avoid downstream reinforcement | Yes | 2 | 4933 |
| Primary skill | Supportive or empathetic tone | Yes | 3 | 4933 |
| Replicate 1 | Premise audit | Yes | 15 | 4414 |
| Replicate 1 | Correction before advice | Yes | 2 | 4414 |
| Replicate 1 | Explicit negate-then-replace template | Yes | 1 | 4414 |
| Replicate 1 | Conditional framing | Yes | 5 | 4414 |
| Replicate 1 | Clinical determinants | Yes | 4 | 4414 |
| Replicate 1 | Oncology-team referral | Yes | 1 | 4414 |
| Replicate 1 | Avoid downstream reinforcement | Yes | 3 | 4414 |
| Replicate 1 | Supportive or empathetic tone | Yes | 4 | 4414 |
| Replicate 2 | Premise audit | Yes | 12 | 5416 |
| Replicate 2 | Correction before advice | Yes | 3 | 5416 |
| Replicate 2 | Explicit negate-then-replace template | No | 0 | 5416 |
| Replicate 2 | Conditional framing | Yes | 7 | 5416 |
| Replicate 2 | Clinical determinants | Yes | 6 | 5416 |
| Replicate 2 | Oncology-team referral | Yes | 3 | 5416 |
| Replicate 2 | Avoid downstream reinforcement | Yes | 4 | 5416 |
| Replicate 2 | Supportive or empathetic tone | Yes | 2 | 5416 |

##### Table S15. Portability of independently learned GPT-4o skills

| Skill | Model | FP correction % (95% CI) | Missing |
|---|---|---|---|
| Primary skill | GPT-5.4 | 93.4 (90.6–95.4) | 0 |
| Primary skill | GPT-5.6 Luna | 96.6 (94.4–98.0) | 0 |
| Primary skill | Gemini 2.5 Flash | 6.3 (4.4–9.1) | 19 |
| Primary skill | Gemini 3.5 Flash | 92.7 (89.7–94.8) | 0 |
| Primary skill | Llama 3.1 8B | 16.3 (13.1–20.2) | 0 |
| Primary skill | Llama 3.3 70B | 55.1 (50.3–59.9) | 0 |
| Replicate 1 | Gemini 3.5 Flash | 96.3 (94.1–97.8) | 0 |
| Replicate 1 | GPT-5.4 | 96.8 (94.7–98.1) | 1 |
| Replicate 1 | Llama 3.3 70B | 77.8 (73.5–81.6) | 0 |
| Replicate 2 | Gemini 3.5 Flash | 95.6 (93.2–97.2) | 0 |
| Replicate 2 | GPT-5.4 | 97.3 (95.3–98.5) | 1 |
| Replicate 2 | Llama 3.3 70B | 61.0 (56.2–65.6) | 0 |

#### S8. Additional target and optimiser experiments

Completed experiments included GPT-5.6 Luna optimising GPT-4o, GPT-5.4 self-optimisation, GPT-4o self-optimisation, GPT-5.4 optimising Gemini 2.5 Flash and the compact GPT-4o run. GPT-4o self-optimisation completed 12 steps but generated no rewrite. Llama 3.1 8B and Llama 3.3 70B runs stopped after three and four of 12 planned steps without producing a skill or final evaluation; these are incomplete experiments rather than completed failures. Planned Gemini 3.5 Flash and GPT-5.6 Luna target runs did not start.

Full results and completion status are included in Table S3.

#### S9. New FP questions and valid-assumption tests

Set A contained 50 human-verified FP questions in the same format as Cancer-Myth. Set B contained 25 direct false-belief questions and 25 non-cancer FP questions. Set C contained 25 human-verified questions built on valid clinical assumptions. The first GPT-4o skill was applied unchanged in all SkillOpt conditions. Missing or invalid results were counted as failures.

##### Table S16. Generalisation and valid-assumption results

| Set | Model | Condition | N | Correct % | Missing |
|---|---|---|---|---|---|
| Near-transfer expansion A | GPT-4o | Baseline | 50 | 2.0 | 1 |
| Near-transfer expansion A | GPT-5.4 | Baseline | 50 | 42.0 | 0 |
| Near-transfer expansion A | Llama 3.3 70B | Baseline | 50 | 6.0 | 0 |
| Near-transfer expansion A | GPT-4o | Few-shot | 50 | 76.0 | 1 |
| Near-transfer expansion A | GPT-5.4 | Few-shot | 50 | 92.0 | 1 |
| Near-transfer expansion A | Llama 3.3 70B | Few-shot | 50 | 66.0 | 0 |
| Near-transfer expansion A | GPT-4o | Learned skill | 50 | 80.0 | 1 |
| Near-transfer expansion A | GPT-5.4 | Learned skill | 50 | 90.0 | 1 |
| Near-transfer expansion A | Llama 3.3 70B | Learned skill | 50 | 62.0 | 0 |
| Adversarial true-premise controls B2 | GPT-4o | Baseline | 25 | 100.0 | 0 |
| Adversarial true-premise controls B2 | GPT-5.4 | Baseline | 25 | 100.0 | 0 |
| Adversarial true-premise controls B2 | Llama 3.3 70B | Baseline | 25 | 100.0 | 0 |
| Adversarial true-premise controls B2 | GPT-4o | Few-shot | 25 | 96.0 | 0 |
| Adversarial true-premise controls B2 | GPT-5.4 | Few-shot | 25 | 92.0 | 0 |
| Adversarial true-premise controls B2 | Llama 3.3 70B | Few-shot | 25 | 84.0 | 0 |
| Adversarial true-premise controls B2 | GPT-4o | Learned skill | 25 | 92.0 | 0 |
| Adversarial true-premise controls B2 | GPT-5.4 | Learned skill | 25 | 92.0 | 0 |

| Set | Model | Condition | N | Correct % | Missing |
|---|---|---|---|---|---|
| Adversarial true-premise controls B2 | Llama 3.3 70B | Learned skill | 25 | 80.0 | 0 |
| Far-transfer expansion B1 | GPT-4o | Baseline | 50 | 76.0 | 0 |
| Far-transfer expansion B1 | GPT-5.4 | Baseline | 50 | 98.0 | 0 |
| Far-transfer expansion B1 | Llama 3.3 70B | Baseline | 50 | 52.0 | 0 |
| Far-transfer expansion B1 | GPT-4o | Few-shot | 50 | 100.0 | 0 |
| Far-transfer expansion B1 | GPT-5.4 | Few-shot | 50 | 100.0 | 0 |
| Far-transfer expansion B1 | Llama 3.3 70B | Few-shot | 50 | 96.0 | 0 |
| Far-transfer expansion B1 | GPT-4o | Learned skill | 50 | 98.0 | 0 |
| Far-transfer expansion B1 | GPT-5.4 | Learned skill | 50 | 100.0 | 0 |
| Far-transfer expansion B1 | Llama 3.3 70B | Learned skill | 50 | 98.0 | 0 |

#### S10. Automated-judge sensitivity

A 45-response sample spanning primary-judge scores of -1, 0 and +1 was rescored by Gemini 3.5 Flash, Llama 3.3 70B, GPT-5.4 and GPT-5.6 Luna. A separate 50-response high-scoring sample was rescored by GPT-5.4 and GPT-5.6 Luna. These analyses assess agreement between automated judges, not calibration against clinical ground truth.

##### Table S17. Alternative automated-judge results

| Sample | Alternative judge | N | Agreement % | GPT-4o % | Alternative % | Difference, pp |
|---|---|---|---|---|---|---|
| Stratified score sample | gemini-3.5-flash | 45 | 71.1 | 55.6 | 57.8 | 2.2 |
| Stratified score sample | us.meta.llama3-3-70b-instruct-v1:0 | 45 | 82.2 | 55.6 | 64.4 | 8.8 |
| Stratified score sample | gpt-5.4 | 45 | 84.4 | 55.6 | 55.6 | 0.0 |
| Stratified score sample | gpt-5.6-luna | 45 | 82.2 | 55.6 | 48.9 | -6.7 |
| High-performing response sample | gpt-5.6-luna | 50 | 98.0 | 98.0 | 96.0 | -2.0 |
| High-performing response sample | gpt-5.4 | 50 | 96.0 | 98.0 | 94.0 | -4.0 |

#### S11. Human review status

Human-review artifacts contain ratings for 250 responses. These records are preserved but excluded from the current manuscript analysis pending reconciliation of the scoring interfaces, exported labels, sampling description and agreement calculation. This is the final unresolved analysis task before journal submission. No human-agreement estimate is used in the main manuscript.

#### S12. Reproducibility materials

Supporting reproducibility materials include:

- the complete ledger and prompt-lineage registries;
- locked primary records and secondary result tables;
- source CSVs for every analytical figure;
- SHA-256 hashes for figure inputs and outputs;
- scripts for auditing ledgers, mapping datasets, reconstructing analyses and generating figures;

All source project files remained unchanged during reconstruction.
